# The transcriptional profile of keloidal Schwann cells

**DOI:** 10.1101/2022.03.16.22272464

**Authors:** Martin Direder, Matthias Wielscher, Tamara Weiss, Maria Laggner, Dragan Copic, Katharina Klas, Daniel Bormann, Vera Vorstandlechner, Erwin Tschachler, Hendrik Jan Ankersmit, Michael Mildner

**Author notes:** **corresponding author:** Michael Mildner, PhD, Department of Dermatology, Medical University of Vienna, Lazarettgasse 14, 1090 Vienna, Austria.

## Abstract

Recently, a specific Schwann cell type with pro-fibrotic and tissue regenerative properties has been identified that contributes to keloid formation. In the present study, we have reanalysed published single cell RNA sequencing (scRNAseq) studies of keloids, healthy skin and normal scars to reliably determine the specific gene expression profile of the keloid-specific Schwann cell type in more detail.

We were able to confirm the presence of the repair-like, pro-fibrotic Schwann cell type in the datasets of all three studies and identified a specific gene set for these Schwann cells. In contrast to keloids, in normal scars the number of Schwann cells was neither increased nor was their gene expression profile distinctly different from Schwann cells of normal skin. In addition, our bioinformatics analysis provided evidence for a role of transcription factors of the kruppel-like factor family and members of the immediate early response genes, in the de-differentiation process of keloidal Schwann cells.

Together, our analysis strengthens the role of the pro-fibrotic Schwann cell type in the formation of keloids. Knowledge on the exact gene expression profile of these Schwann cells will facilitate their identification in other organs and diseases.

## Introduction

Schwann cells are glia cells of the peripheral nervous system and ensure proper nerve development and integrity ^1^. After peripheral nerve injury, mature Schwann cells undergo transcriptional reprogramming that involves de-differentiation into an immature cell state and the acquisition of repair specific functions ^2-8^. These repair Schwann cells are essential to orchestrate nerve regeneration by attracting immune cells to the site of injury, phagocytosis of myelin debris, secretion of neurotrophic and neuritogenic factors and formation of regeneration tracks (Bungner bands) to stimulate and guide re-growing axons ^9-14^. De-differentiation into repair Schwann cells has been shown to involve the expression and activation of a variety of specific factors ^15^, including Jun proto-oncogene (*JUN*), signal transducer and activator of transcription 3 (*STAT3*), brain-derived neurotrophic factor (*BDNF*), artemin (*ARTN*), insulin like growth factor binding protein 2 (*IGFBP2*) and glial cell line-derived neurotrophic factor (*GDNF*) ^16-20^. Especially, oligodendrocyte transcription factor 1 (*OLIG1*) and sonic hedgehog (*SHH*) have been suggested as specific markers for repair Schwann cells ^9^.

In healthy skin, Schwann cells persist in myelinating and non-myelinating states ^1^. Recently, Parfejevs et al. provided evidence that Schwann cells not only contribute to the regeneration of the nerve, but also to dermal wound healing by proliferating and emanating from the disrupted nerve. These skin-derived repair Schwann cells populate the damaged area, where they support the differentiation of fibroblasts into myofibroblasts, promote wound contraction and closure as well as re-epithelization ^21^. After completion of neuronal regeneration and wound healing, skin repair Schwann cells re-differentiate into their adult state and ensheath the restored axons ^10^. Ideally, the entire healing process results in an asymptomatic, fine lined, flat scar.

Abnormal scars, such as hypertrophic scars or keloids, can result in serious health problems, including movement restrictions, persistent itch and pain ^22-25^. Keloids represent a special type of scars, characterised by a tumour-like continuous growth beyond the margins of the original wound ^22^. They exclusively develop in humans and the lack of adequate model systems complicates basic research into their pathogenesis ^26,27^. Therefore, in spite of several decades of research, the exact mechanistic events driving keloid formation remain largely unclear ^28^.

The recent development of single cell RNA sequencing (scRNAseq) enables a completely new approach to decode disease pathomechanisms. So far, three research groups have applied scRNAseq to study keloid tissue on a cellular and transcriptional level. Liu *et al*. compared the centre of keloids with adjacent skin. They identified various dysregulated genes and pathways in keloidal fibroblasts and endothelial cells and unravelled TWIST1 as an important factor in keloidal fibrogenesis ^29^. Deng *et al*. applied scRNAseq to compare keloids with normal scars ^30^. They subdivided the keloidal fibroblasts into four major groups and identified an increase of extracellular matrix (ECM)-producing mesenchymal fibroblasts in keloid tissue. Whereas these studies focused on fibroblasts and endothelial cells, a recent study by our group demonstrated that keloids contain an increased number of phenotypically distinct Schwann cells ^31^. The vast majority of these keloidal Schwann cells displayed a cellular state comparable to the one described for Schwann cells in regenerating nerves ^31^. Our additional finding that these Schwann cells expressed multiple genes associated with matrix formation makes them highly plausible candidates for playing a crucial role in the developments of keloids ^31^.

Due to donor variabilities, differences in sample preparation and data processing, scRNAseq studies are not always straightforwardly comparable ^32-35^. To obtain more consistent data, individual datasets of different laboratories should be re-evaluated in a combined analysis, as previously shown for skin fibroblast populations ^36^. This study clearly demonstrated that, despite differences in the experimental procedures, major similarities amongst all fibroblast populations are conserved across the scRNAseq data sets published by the different laboratories. Studies on the comparability of smaller cell clusters, such as Schwann cells, in scRNAseq datasets are so far not available.

Here, we performed a comparative analysis of Schwann cells in different scRNAseq datasets of healthy skin, normal scars, keloids, and keloid adjacent skin from four independent research groups to elucidate the gene set most reliably identifying the keloidal Schwann cell population. In addition, we compared two distinct bioinformatics approaches for the analysis of numerous different scRNAseq data sets revealing significant variations of the two calculation methods. Our study shows that the presence of the previously described repair-like, pro-fibrotic Schwann cells in keloids is indeed conserved across different scRNAseq data sets and provides a specific expression pattern for keloidal Schwann cells. This so far unrecognized cells might represent a novel, potential target for keloid treatment.

## Materials and Methods

### Data acquisition

In this study, scRNAseq-datasets of 6 healthy skin-, 6 normal scar-, 11 keloid- and 4 keloid adjacent skin-samples were analysed (Figure S1a). ScRNAseq data of normal scars and keloids have already been generated by Direder *et al*. and are deposited in NCBI’s Gene Expression Omnibus (GSE181316) ^31^. Skin data published by Tabib *et al*. (2018) were downloaded from https://dom.pitt.edu/wp-content/uploads/2018/10/Skin_6Control_rawUMI.zip and https://dom.pitt.edu/wp-content/uploads/2018/10/Skin_6Control_Metadata.zip ^32^. ScRNAseq data of keloids and normal scars generated by Deng *et al*. were downloaded from the Gene Expression Omnibus database (dataset GSE163973) ^30^. Transcriptomic data from Liu *et al*. were downloaded from the Genome Sequencing Archive (BioProject PRJCA003143) ^29^. Count tables were obtained from the original fastq read files applying the cellranger pipeline (10X Genomics CellRanger 3.0.2, Pleasonton, CA, USA) ^37^.

### Data processing

Computational analyses were performed utilizing R and R-studio (version 4.0.3, The R Foundation, Vienna, Austria). All features of the included datasets were screened, and doublets were removed. The feature designation was harmonized and only features detected in all datasets were included in subsequent analyses. Datasets were transformed into a Seurat object using Seurat (Seurat v4.0.1, Satija Lab) ^38^. The sctransform-normalization in combination with the glmGamPoi package and removal of mitochondrial mapping percentage were applied to preprocess the data ^39,40^. Erythrocytes were excluded by removing cells with a Hemoglobin Subunit Beta (HBB) expression >5. For this study, the included datasets were combined all together and individually by source and tissue type (Figure S1b). All dataset combinations were processed according to the Seurat Vignette. Principal Component Analysis (PCA), Uniform Manifold Approximation (UMAP) and Projection dimensionality reduction were applied according to the Seurat protocol. Clusters were identified as cell types according to their expression of well-established marker genes ^31^. Schwann cell clusters were subsetted and data were preprocessed and calculated in the same way as described above. The commands “RunUMAP” and “FindNeighbors” were performed on 30 dimensions, “FindClusters” was applied with a resolution of “0.3”. Subset clusters expressing marker genes typical for melanocytes were removed. Schwann cells were characterized by the expression of established marker genes ^31,41^. Differentially expressed genes were defined as genes displaying an average fold change > 2. The SC-Keloid-specific gene expression pattern was identified by examination of SC-Keloid against all myelinating and nonmyelinating Schwann cells of all dataset combinations (Tables S1-5). From these lists, the top 100 upregulated genes were compared and only genes included in all gene lists were selected as members of the SC-Keloid gene expression pattern. Genes without a clear demarcation from the myelinating and non-myelinating Schwann cells were excluded. Gene ontology enrichment analysis was performed using Metascape with 0.05 as p-value cutoff and 2 as minimum enrichment score ^42^. Only corresponding GOs with “Summary” GroupID were shown. For pseudotime analysis, the Schwann cell cluster from all single calculated datasets were combined and preprocessed the same way. As normal scar datasets from both sources revealed no Schwann cell cluster, these datasets were excluded from pseudotime analysis. The commands “RunUMAP” and “FindNeighbors” were performed on 18 dimensions, “FindClusters” was applied with a resolution of 0.5. Monocle3 (Monocle3, v.0.2.3.0, Trapnell Lab, Seattle, Washington, USA) was used to create the pseudotime trajectory ^43-47^. The principal graph was not pruned and a total of 7 centers were determined. The generated metascape lists were matched and only –log10(P) values of terms identified in all lists were pictured together with their arithmetic mean. To identify upregulated genes specific for the branching point in the combined Schwann cell object, the command “Findmarkers” was executed, comparing the newly identified Seurat_cluster located in the branching point against all remaining Schwann cells.

### Sequencing data juxtaposition to gene expression profile of peripheral nerve regeneration

The human homologs of the gene list published by Bosse *et al*. ^15^ were identified using the GeneBank accession number and the website Uniprot (https://www.uniprot.org/, accessed on 2021-10-18). Relative expression levels were identified using the command “FindMarkers” with SC-Keloid as ident.1 for up regulation and 2 for down regulation, the myelinationg and nonmyelinating cluster as the opposite ident, a min.pct of 0.01 and a logfc.threshold of 0.01 were set. Expression changes < 0.01 were set as 0.

### Collagen alignment examination

Hematoxylin and eosin stainings of healthy skin, normal scar, and keloid samples were imaged. Alignment of collagen bundles were determined applying Curvealign V4.0 Beta (MATLAB software, Cleve Moler, MathWorks, Natick, Massachusetts, USA). Collagen color, contrast and brightness were edited by Adobe Photoshop CS6 (Adobe Inc, San Jose, CA, USA). Region of interest (ROI) was defined by size (256 height, 256 width) and four ROIs per condition were analyzed. The coefficiency of alignment was statistically evaluated.

### Immunofluorescence

Tissue samples were fixed in neutral buffered 4.5% formaldehyde (SAV Liquid Production GmbH, Flintsbach am Inn, Germany) at 4°C overnight, followed by a washing step with phosphate-buffered saline (PBS) overnight. Dehydration was performed by sequential incubations with 10%, 25% and 42% sucrose for 24 hours each. Tissues were embedded in optimal cutting temperature compound (OCT compound, TissueTek, Sakura, Alphen aan den Rijn, The Netherlands) and stored at -80°C. Samples were cut into 10 µm-thick sections and dried for 30 min at room temperature. Slides were blocked and permeabilized for 15 min with 1% BSA, 5% goat serum (DAKO, Glostrup, Denmark), and 0.3% Triton-X (Sigma Aldrich, St. Louis, MO, USA) in 1xPBS. Sections were incubated with S100 (rabbit anti-human, ready to use, DAKO) primary antibody solution overnight at 4°C followed by three washing steps with 1x PBS for 5 min each. Sections were incubated with secondary antibody (goat anti-rabbit, alexa fluor 488-conjugated, 1:600, Invitrogen, Waltham, MA, USA) and 50 µg/ml 4,6-diamidino-2-phenylindole (DAPI, Thermo Fisher Scientific, Waltham, MA, USA) in 1xPBS for 50 min at room temperature. After three washing steps, sections were covered using mounting medium and stored at 4°C. Images were acquired using a confocal laser scanning microscope (TCS SP8X, Leica, Wetzlar, Germany) equipped with a 10x (0.3 HCPL FluoTar), a 20x (0.75 HC-Plan-Apochromat, Multimmersion), a 20x (0.75 HC-Plan-Apochromat) and a 63x (1.3 HC-Plan-Apochromat, Glycerol) objective using Leica application suite X version 1.8.1.13759 or LAS AF Lite software (both Leica). Final images constitute a maximum projection of total z-stacks.

### Statistics

Statistical evaluation was performed using GraphPad Prism 8 software (GraphPad Software Inc., La Jolla, CA, USA). Shapiro-Wilk test was used to test for normal distribution. One-way ANOVA with Tukey *post hoc* test was used to compare three and more groups. P-values were marked with asterisks: *p<0.05, **p<0.01, ***p<0.001 ****p<0.0001.

## Results

### Combined integration of all scRNAseq data sets identifies significant differences in the cellular composition of normal skin, normal scars and keloids

Histological examination of normal skin samples, normal scars and keloids showed increased dermal cell numbers and abnormalities of the ECM in both types of scars, including increased linearity of collagen bundles (Figure 1a and Figure S2) ^48^. To determine how cell populations differ between healthy skin, skin adjacent to keloids, normal scars and keloids, we performed a combined integration of individual scRNAseq data sets ^29-32^. In total, datasets of six normal skin samples, four skin samples adjacent to keloids, six normal scars and eleven keloids were included. Demographic data on the samples and methodological differences in sample preparation are shown in Tables 1 and 2, respectively. Combined bioinformatics analysis of the datasets revealed several main cell clusters (Figure 1b), which were further characterized by marker gene expression (Figure 1c). The clusters were identified as fibroblasts (FB), smooth muscle cells (SMC), pericytes (PC), keratinocytes (KC), endothelial cells (EC), lymphatic endothelial cells (LEC), T-cells (TC), macrophages (MAC), dendritic cells (DC), Schwann cells (SC) and melanocytes (MEL) (Figure 1b and c).

**Table 1.** donor information

| Tissue | Source | Paper ID | Original ID | Site | Age | Sex | Race |
| --- | --- | --- | --- | --- | --- | --- | --- |
| normal skin | TT<sup>1</sup> | TT1_s | SC18control | forearm | 63 | male | caucasian |
| normal skin | TT<sup>1</sup> | TT2_s | SC1control | forearm | 54 | male | caucasian |
| normal skin | TT<sup>1</sup> | TT3_s | SC32control | forearm | 66 | female | caucasian |
| normal skin | TT<sup>1</sup> | TT4_s | SC33control | forearm | 23 | female | asian |
| normal skin | TT<sup>1</sup> | TT5_s | SC34control | forearm | 62 | female | caucasian |
| normal skin | TT<sup>1</sup> | TT6_s | SC4control | forearm | 24 | male | caucasian |
| adjacent skin | XL<sup>2</sup> | XL1_s | K007CTRL | chest | 28 | female | chinese |
| adjacent skin | XL<sup>2</sup> | XL2_s | K009CTRL | chest | 32 | female | chinese |
| adjacent skin | XL<sup>2</sup> | XL3_s | K013CTRL | chest | 26 | female | chinese |
| adjacent skin | XL<sup>2</sup> | XL4_s | K012CTRL | chest | 26 | female | chinese |
| normal scar | CCD<sup>3</sup> | CCD1_ns | human_normal_scar_sample1 | back | 39 | male | han nationality |
| normal scar | CCD<sup>3</sup> | CCD2_ns | human_normal_scar_sample2 | chest | 28 | male | han nationality |
| normal scar | CCD<sup>3</sup> | CCD3_ns | human_normal_scar_sample3 | chest | 26 | female | han nationality |
| normal scar | MD<sup>4</sup> | MD1_ns | scar1 | abdomen | 26 | female | caucasian |
| normal scar | MD<sup>4</sup> | MD2_ns | scar2 | abdomen | 43 | female | caucasian |
| normal scar | MD<sup>4</sup> | MD3_ns | scar3 | abdomen | 60 | male | caucasian |
| keloid | XL<sup>2</sup> | XL1_k | K007CASE | chest | 28 | female | chinese |
| keloid | XL<sup>2</sup> | XL2_k | K009CASE | chest | 32 | female | chinese |
| keloid | XL<sup>2</sup> | XL3_k | K013CASE | chest | 26 | female | chinese |
| keloid | XL<sup>2</sup> | XL4_k | K012CASE | chest | 26 | female | chinese |
| keloid | CCD<sup>3</sup> | CCD1_k | human_keloid_sample1 | back | 20 | male | han nationality |
| keloid | CCD<sup>3</sup> | CCD2_k | human_keloid_sample2 | chest | 23 | male | han nationality |
| keloid | CCD<sup>3</sup> | CCD3_k | human_keloid_sample3 | chest | 34 | female | han nationality |
| keloid | MD<sup>4</sup> | MD1_k | keloid_1 | chest | 36 | female | caucasian |
| keloid | MD<sup>4</sup> | MD2_k | keloid_2 | earlobe | 60 | female | african |
| keloid | MD<sup>4</sup> | MD3_k | keloid_3L | earlobe - left | 34 | male | caucasian |
| keloid | MD<sup>4</sup> | MD4_k | keloid_3R | earlobe - right | 34 | male | caucasian |

1. Tabib, T.; Morse, C.; Wang, T.; Chen, W.; Lafyatis, R. SFRP2/DPP4 and FMO1/LSP1 Define Major Fibroblast Populations in Human Skin. *J Invest Dermatol* 2018, 138, 802-810. 2. Liu, X.; Chen, W.; Zeng, Q.; Ma, B.; Li, Z.; Meng, T.; Chen, J.; Yu, N.; Zhou, Z.; Long, X. Single-cell RNA-seq reveals lineage-specific regulatory changes of fibroblasts and vascular endothelial cells in keloids. *J Invest Dermatol* 2021. 3. Deng, C.C.; Hu, Y.F.; Zhu, D.H.; Cheng, Q.; Gu, J.J.; Feng, Q.L.; Zhang, L.X.; Xu, Y.P.; Wang, D.; Rong, Z.; et al. Single-cell RNA-seq reveals fibroblast heterogeneity and increased mesenchymal fibroblasts in human fibrotic skin diseases. *Nat Commun* 2021, 12, 3709. 4. Direder, M.; Weiss, T.; Copic, D.; Vorstandlechner, V.; Laggner, M.; Mildner, C.S.; Klas, K.; Bormann, D.; Haslik, W.; Radtke, C.; et al. Schwann cells contribute to keloid formation. *medRxiv* 2021, 2021.2008.2009.21261701.

**Table 2.**
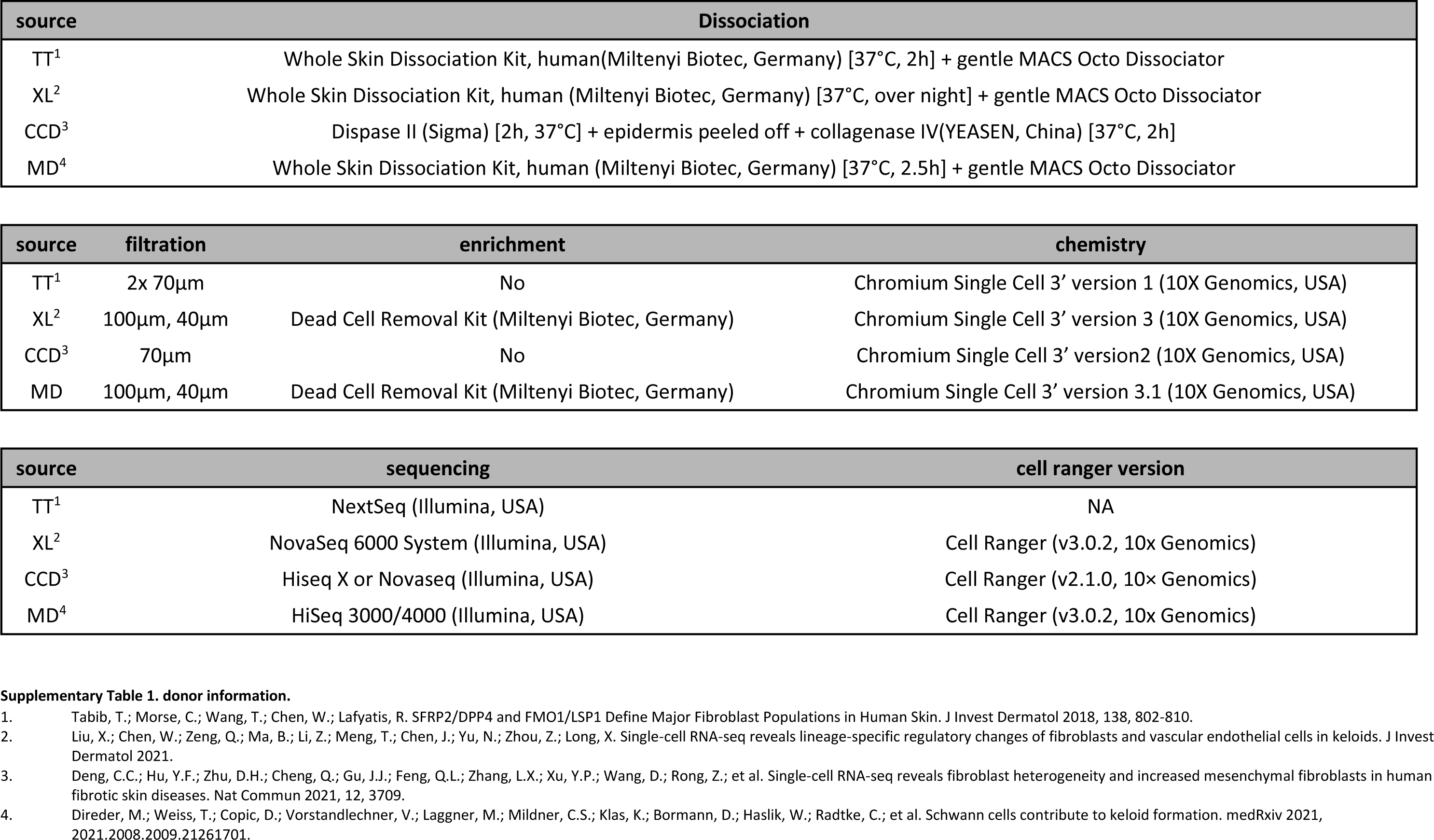
comparison_methods & materials

**Figure 1:**
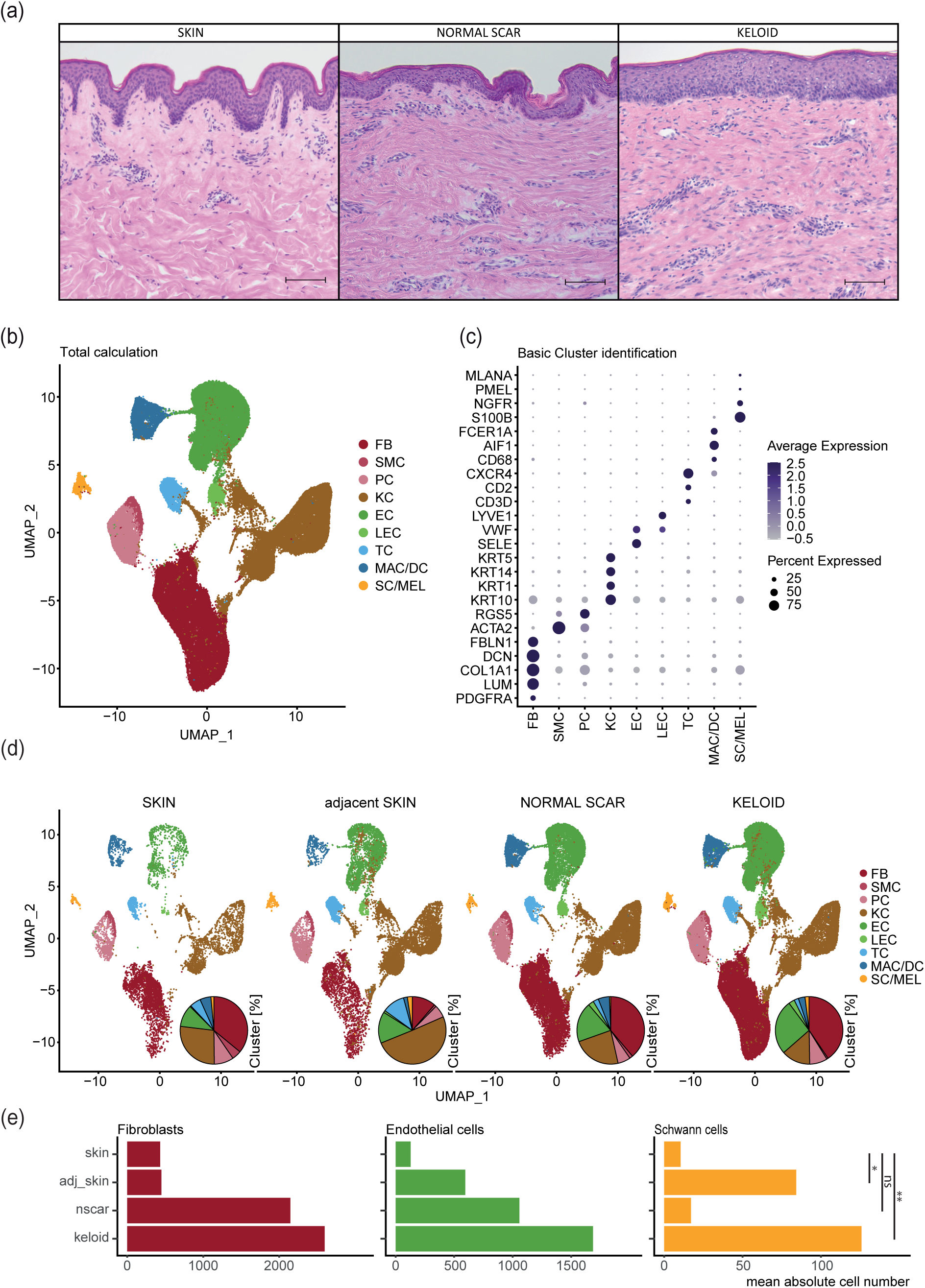
Cellular composition of skin, normal scars, keloids and keloid-adjacent skin. (a) Hematoxylin-Eosin stainings of skin, normal scar and keloid. Scale bars: 250 µm. (b) UMAP-Plot after integration of all datasets. Cluster identifications of fibroblasts (FB), smooth muscle cells (SMC), pericytes (PC), keratinocytes (KC), endothelial cells (EC), lymphatic endothelial cells (LEC), T-cells (TC), macrophages and dendritic cells (MAC/DC), Schwann cells and melanocytes (SC/MEL). (c) Dotplots showing well-known marker genes to characterise clusters: platelet-derived growth factor receptor A (*PDGFRA*), lumican (*LUM*), collagen type I alpha 1 (*COL1A1*), decorin (*DCN*), fibulin-1 (*FBLN1*) for FB, actin alpha 2 (*ACTA2*), regulator of G protein signalling 5 (*RGS5*) for SMC and PC, keratin 10 (*KRT10*), keratin 1 (*KRT1*), keratin 14 (*KRT14*), keratin 5 (*KRT5*) for KC, E-selectin (*SELE*), von willebrand factor (*VWF*) for EC, lymphatic vessel endothelial hyaluronan receptor 1 (*LYVE1*) for LEC, *CD3D*, cluster of differentiation 2 (*CD2*), c-x-c chemokine receptor type 4 (*CXCR4*) for TC, cluster of differentiation 68 (*CD68*), allograft inflammatory factor 1 (*AIF1*) for MAC, Fc fragment of IgE receptor Ia (*FCER1A*) for DC, S100 calcium binding protein B (*S100B*), nerve growth factor receptor (*NGFR*) for SC, premelanosome protein (*PMEL*), melan-a (*MLANA*) for MEL; Dot size symbolizes percentage of cells expressing the gene, colour gradient represents average gene expression. (d) Split UMAP-Plots show cellular composition within each tissue. Pie plots depict relative amounts of each cell type within a tissue. (e) Bar plots depict arithmetic means of the absolute numbers of FB, EC and SC within each condition. Asterisks represent p-values: *p<0.05, **p<0.01, ***p<0.001, ns: not significant:

Interestingly, macrophages and dendritic cells as well as Schwann cells and melanocytes clustered together, respectively (Figure 1b). Split analysis revealed similar cell clusters but different cell numbers in each condition (Figure 1d). While cell numbers of fibroblast and endothelial cells were increased in normal scars and keloids, increased numbers of Schwann cells were only detectable in keloids, indicating a specific role of Schwann cells in the pathogenesis of keloids (Figure 1e).

### Keloidal Schwann cells are conserved in different scRNAseq data sets

As the main goals of our study were the in-depth analysis of Schwann cells in the different scar types and their comparability across different single cell data sets, we next recalculated the combined integrated Schwann cell cluster, removed all melanocytes and performed subclustering (Figure 2). The different Schwann cell subclusters (Figure 2a) were identified using well-established Schwann cell markers (Figure 2b) ^31,41^. This analysis identified myelinating Schwann cells (SC-Myel), nonmyelinating Schwann cells (SC-Nonmyel), proliferating Schwann cells (SC-Prolif), Schwann cells additionally expressing genes typical for endothelial cells (SC-EC) or fibroblasts (SC-FB) and the previously described repair-like, pro-fibrotic keloidal Schwann cells (SC-Keloid) (Figure 2a and b). We next separated each dataset and detected only few Schwann cells in normal skin (Figure 2c). In contrast, the amount of Schwann cells was remarkably increased in keloids (Figure 2c). Interestingly, we also found an increased number of Schwann cells in skin adjacent to keloids but not in normal scars (Figure 2c). Schwann cells of normal skin were identified as either myelinating or non-myelinating Schwann cells. By contrast, we found a high plasticity of Schwann cells in keloids, with a high number of cells displaying a pro-fibrotic phenotype. These keloidal Schwann cells were also detected in one skin sample adjacent to keloids (Figure 2c). Of note, a substantial number of cells in the Schwann cell cluster present in normal scars did not express the Schwann cell marker gene *S100B* but genes typical for fibroblasts, such as lumican (*LUM*), suggesting wrong cluster assignment of some fibroblasts in the combined integration (Figure 2d, single red dots in normal scar). Contrary, some keloids contained Schwann cells co-expressing fibroblasts markers and *S100B* (Figure 2d, double positive, yellow dots in keloids). To confirm our transcriptomic data, we performed S100 immuno-stainings of normal skin, normal scars and keloids (Figure 2e). As suggested by our scRNAseq data, we found a markedly increased number of Schwann cells in keloids. These cells were spindle shaped with long extensions on both ends (Figure 2e), a morphology previously described for repair Schwann cells ^10,31,49^.

**Figure 2:**
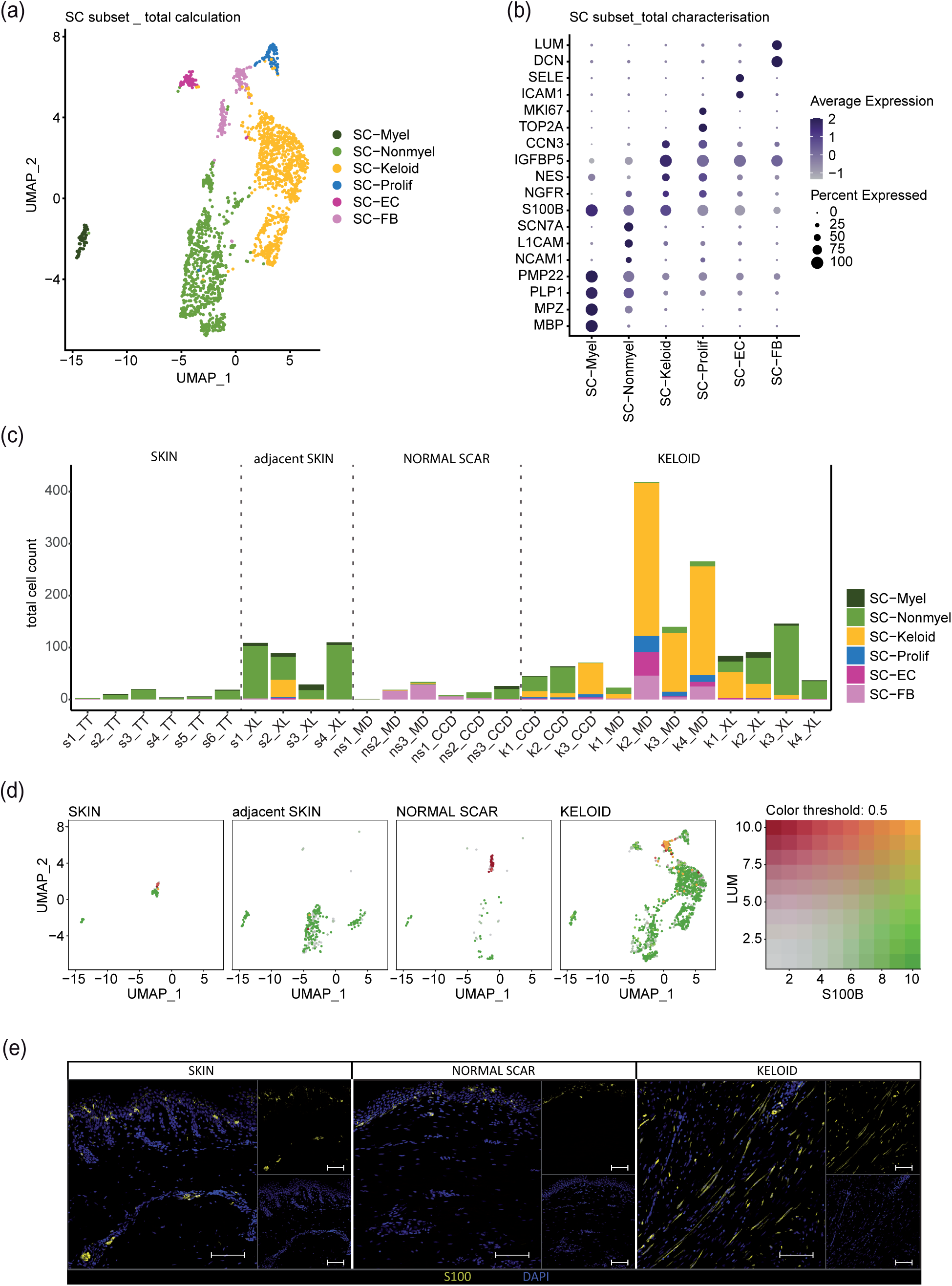
Schwann cell subset detects pro-fibrotic Schwann cells in datasets of all sources. (a) UMAP-Plot of Schwann cell subset (SC subset) after melanocyte removal. (b) Dot plot identifying the Schwann cell subcluster by expression of marker genes: myelin basic protein (*MBP*), myelin protein zero (*MPZ*), proteolipid protein 1 (*PLP1*), peripheral myelin protein 22 (*PMP22*) for myelinating Schwann cells (SC-Myel); neural cell adhesion molecule 1 (*NCAM1*), L1 cell adhesion molecule (*L1CAM*), sodium channel protein type 7 (*SCN7A*) for nonmyelinating Schwann cells (SC-Nonmyel); S100 calcium binding protein B (*S100B*), nerve growth factor receptor (*NGFR*) as general Schwann cell marker; nestin (*NES*), insulin-like growth factor binding protein 5 (*IGFBP5*), cellular communication network factor 3 (*CCN3*) for keloidal Schwann cells (SC-Keloid); DNA topoisomerase II alpha (*TOP2A*), marker of proliferation Ki-67 (*MKI67*) for proliferating Schwann cells (SC-Prolif), decorin (*DCN*), lumican (*LUM*) for cells expressing Schwann cell and fibroblast specific genes (SC-FB), intercellular adhesion molecule 1 (*ICAM1*), E-selectin (*SELE*) for cells expressing Schwann cell and endothelial cell specific genes (SC-EC). (c) Barplots depicting the absolute amount of distinct Schwann cell subtypes within each dataset. skin (s), normal scar (ns), keloid (k); data source: Tracy Tabib et al., 2018 (TT); Xuanyu Liu et al., 2021 (XL), Cheng-Cheng Deng et al., 2021 (CCD), Martin Direder et al., 2021 (MD); (d) Featureblends show the expression of *S100B* (green), *LUM* (red) and double expression of both genes (yellow) in the SC subset split by tissue. (e) Immunofluorescence staining of S100-positive SCs in the dermal layer of a keloid; Scale bar: 100 µm.

To overcome possible calculation errors of combined integration, we also integrated the data sets of each study individually. In line with our previous calculation approach, we found increased numbers of Schwann cells in keloids (Figure S3 und S4). Interestingly, we were not able to identify a Schwann cell cluster in normal scars by separate integration of the data sets (Figure S3). Therefore, only healthy skin, adjacent skin and the three keloid datasets were included for further analysis. All main clusters and subclusters were calculated using the same R-protocol with corresponding identical parameters. Subset analysis uncovered up to five clusters of Schwann cell subtypes in the individual conditions (Figure 3a). Similar to the combined calculation, individual integration of each data set also identified the presence of pro-fibrotic Schwann cells in almost all keloids and in one skin sample adjacent to a keloid (Figure 3a and b). In contrast to the combined integration, we detected a high number of *S100B* negative cells which were lumican-positive in the single integrated skin samples (Figure 3c, single positive red dots). Together, our data indicate that pro-fibrotic Schwann cells are robustly detectable in keloids irrespective of the calculation method. Problems with wrongly assigned fibroblasts are found in both calculation methods and mainly affect very small Schwann cell clusters as present in normal skin or normal scars (Figure 3d).

**Figure 3:**
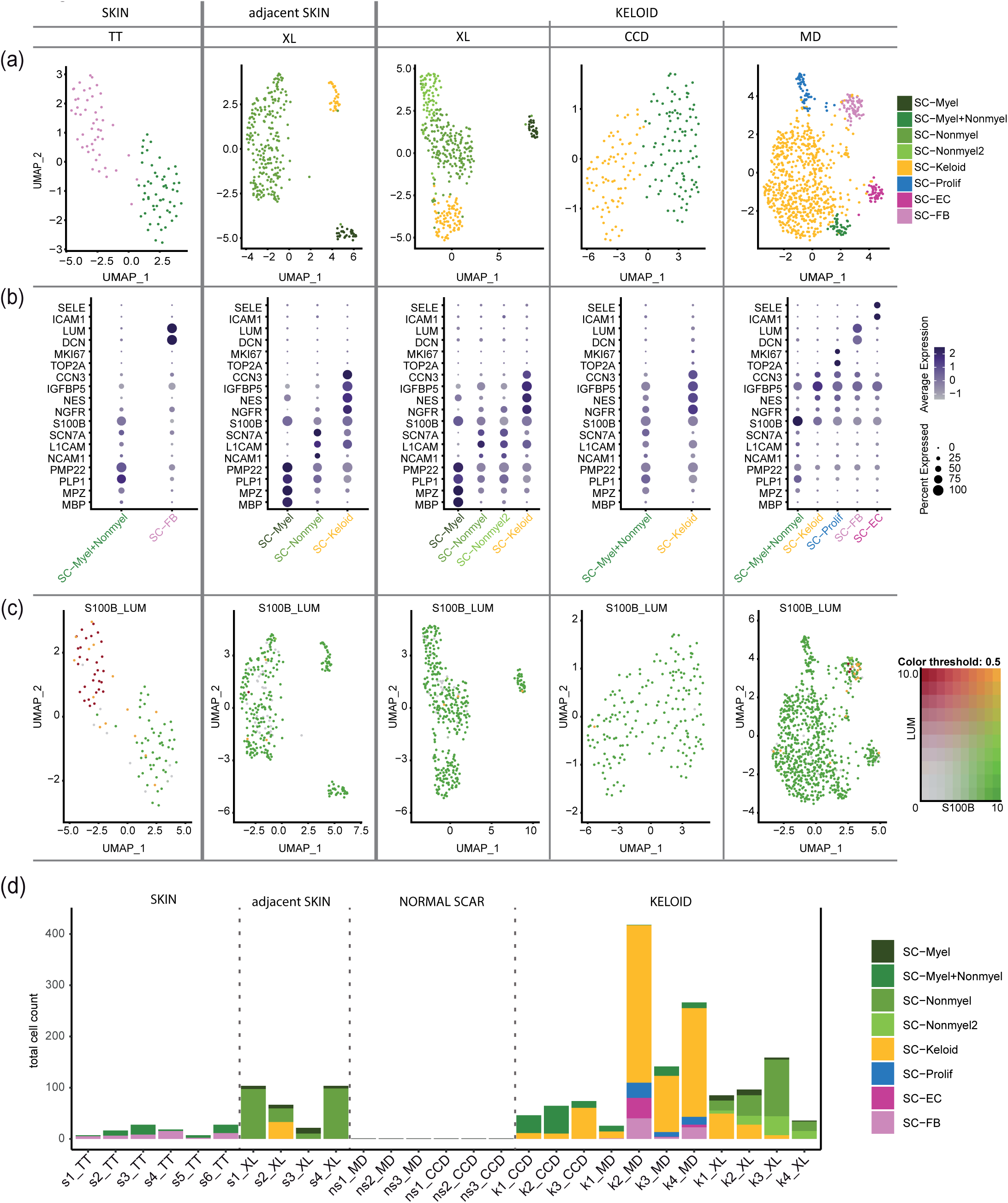
Tissue- and source-specific dataset evaluation confirms keloidal pro-fibrotic Schwann cells. (a) UMAP-Plots depict subsets of identified Schwann cell clusters. Data sources: Tracy Tabib *et al*., 2018 (TT); Xuanyu Liu *et al*., 2021 (XL), Cheng-Cheng Deng *et al*., 2021 (CCD), Martin Direder *et al*., 2021 (MD); (b) Dotplots of well-known marker marker genes to characterize Schwann cell subtypes. myelin basic protein (*MBP*), myelin protein zero (*MPZ*), proteolipid protein 1 (*PLP1*), peripheral myelin protein 22 (*PMP22*) for myelinating Schwann cells (SC-Myel); neural cell adhesion molecule 1 (*NCAM1*), L1 cell adhesion molecule (*L1CAM*), sodium channel protein type 7 (*SCN7A*) for nonmyelinating Schwann cells (SC-Nonmyel, SC-Nonmyel2); S100 calcium binding protein B (*S100B*), nerve growth factor receptor (*NGFR*) as general Schwann cell marker; nestin (*NES*), insulin-like growth factor binding protein 5 (*IGFBP5*), cellular communication network factor 3 (*CCN3*) for keloidal Schwann cells (SC-Keloid); DNA topoisomerase II alpha (*TOP2A*), marker of proliferation Ki-67 (*MKI67*) for proliferating Schwann cells (SC-Prolif), decorin (*DCN*), lumican (LUM) for cells expressing Schwann cell and fibroblast specific genes (SC-FB), intercellular adhesion molecule 1 (*ICAM1*), E-selectin (*SELE*) for cells expressing Schwann cell and endothelial cell specific genes (SC-EC); mixed myelinating and nonmyelinating Schwann cell cluster (SC-Myel+Nonmyel); Colour codes indicate average gene expression levels; Dot sizes visualize relative amounts of positive cells. (c) Feature blends unravel cells positive for *S100B* (green), *LUM* and double positive cells (yellow). (d) Barplots show absolute cell count of distinct Schwann cell cluster in each included dataset. skin (s), normal scar (ns), keloid (k); data source: Tracy Tabib et al., 2018 (TT); Xuanyu Liu et al., 2021 (XL), Cheng-Cheng Deng et al., 2021 (CCD), Martin Direder et al., 2021 (MD).

### Characterization of a gene set specific for keloidal, pro-fibrotic Schwann cells

We next analysed all differentially expressed genes by comparing one Schwann cell cluster with all other Schwann cells within each dataset. Interestingly, more genes were down-regulated in pro-fibrotic Schwann cells than upregulated (Figure S5, Tables S1-5). Evaluation of the most differentially expressed genes between pro-fibrotic Schwann cells and myelinating/non-myelinating Schwann cells revealed a set of 21 genes which were characteristic for the pro-fibrotic Schwann cells in keloids (Figure 4a and b). Comparison of the mean expression values of these genes in the individual data sets confirmed the highly specific expression pattern of keloidal Schwann cells (Figure 4a). Interestingly, expression of most of these genes was comparable between some of the keloid-specific Schwann cell clusters (SC-Prolif, SC-EC and SC-FB) and the pro-fibrotic Schwann cells (SC-Keloid), indicating that although SC-Prolif, SC-EC and SC-FB cluster separately and show some additional characteristics, they also share most of the features of keloidal pro-fibrotic Schwann cells (Figure S6). In our previous work, we discovered a contribution of Schwann cells to ECM formation in keloids ^31^. We therefore analysed expression of matrix proteins in all Schwann cells of the different data sets and found that most collagen genes were significantly increased in keloidal Schwann cells in all data sets (Figure S7). Furthermore, GO-term analysis of keloidal Schwann cell in all datasets confirmed a strong association of these cells with ECM organization and wounding healing (Figure S8).

**Figure 4:**
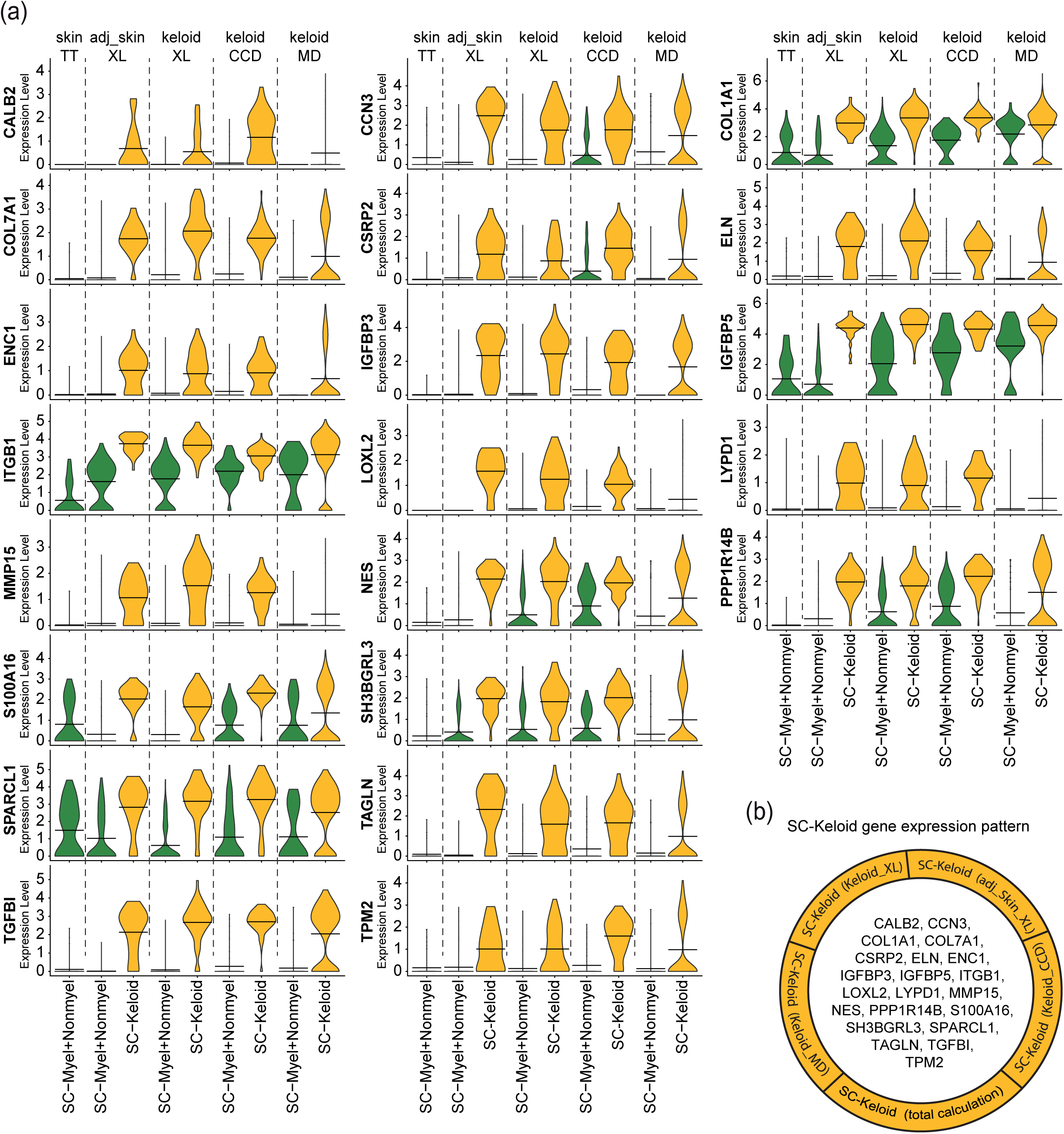
Individual dataset combinations reveal gene expression pattern of keloidal pro-fibrotic Schwann cells. (a) Violin Plots of corresponding, meaningful genes detected in all individual top 100 gene lists comparing keloidal Schwann cells (SC-Keloid) with mature Schwann cells (myelinating and nonmyelinating Schwann cells). Data sources: Tracy Tabib *et al*., 2018 (TT); Xuanyu Liu *et al*., 2021 (XL), Cheng-Cheng Deng *et al*., 2021 (CCD), Martin Direder *et al*., 2021 (MD); adjacent skin (adj_skin); Crossbeams show mean expression values; maximum expression is depicted by vertical lines; frequency of cells with the respective expression level is shown by violin width; myelinating Schwann cells (SC-Myel); nonmyelinating Schwann cells (SC-Nonmyel, SC-Nonmyel2), myelination and nonmyelinating Schwann cell mixed cluster (SC-Myel+Nonmyel); keloidal Schwann cells (SC-Keloid); proliferating Schwann cells (SC-Prolif), cells expressing Schwann cell and endothelial cell specific genes (SC-EC); cells expressing Schwann cell and fibroblast specific genes (SC-FB). (b) List of all members from the SC-Keloid gene expression pattern.

### Pseudotime trajectory analysis identifies candidate factors decisive for keloidal, pro-fibrotic Schwann cell development

Bosse *et al*. were the first to describe a specific panel of genes regulated in nerve-associated cells upon damage of peripheral nerves ^15^. This set of genes was associated with the formation of repair Schwann cells and their function to promote regeneration of the peripheral nerve ^49^. To investigate similarities of repair Schwann cells, present in the injured nerve, and keloidal Schwann cells, we compared the gene list of Bosse *et al*. with our data set and detected 40 genes which were similarly regulated in both data sets (Figure 5). For example, actin beta (*ACTB*), glypican 1 (*GPC1*), myosin heavy chain 9 (*MYH9*), S100 calcium binding protein A4 (*S100A4*), transforming growth factor beta induced (*TGFBI*), ATPase Na+/K+ transporting subunit alpha 2 (*ATP1A2*), myelin and lymphocyte protein (*MAL*), myelin protein zero (*MPZ*), NFKB inhibitor alpha (*NFKBIA*), plasmolipin (*PLLP*), proteolipid protein 1 (*PLP1*) and sodium voltage-gated channel alpha subunit 7 (*SCN7A*) showed comparable regulation (Figure 5, Table S6). These data suggest that a part of the repair Schwann cell gene set mentioned by Bosse *et al*. is conserved in keloidal Schwann cells.

**Figure 5:**
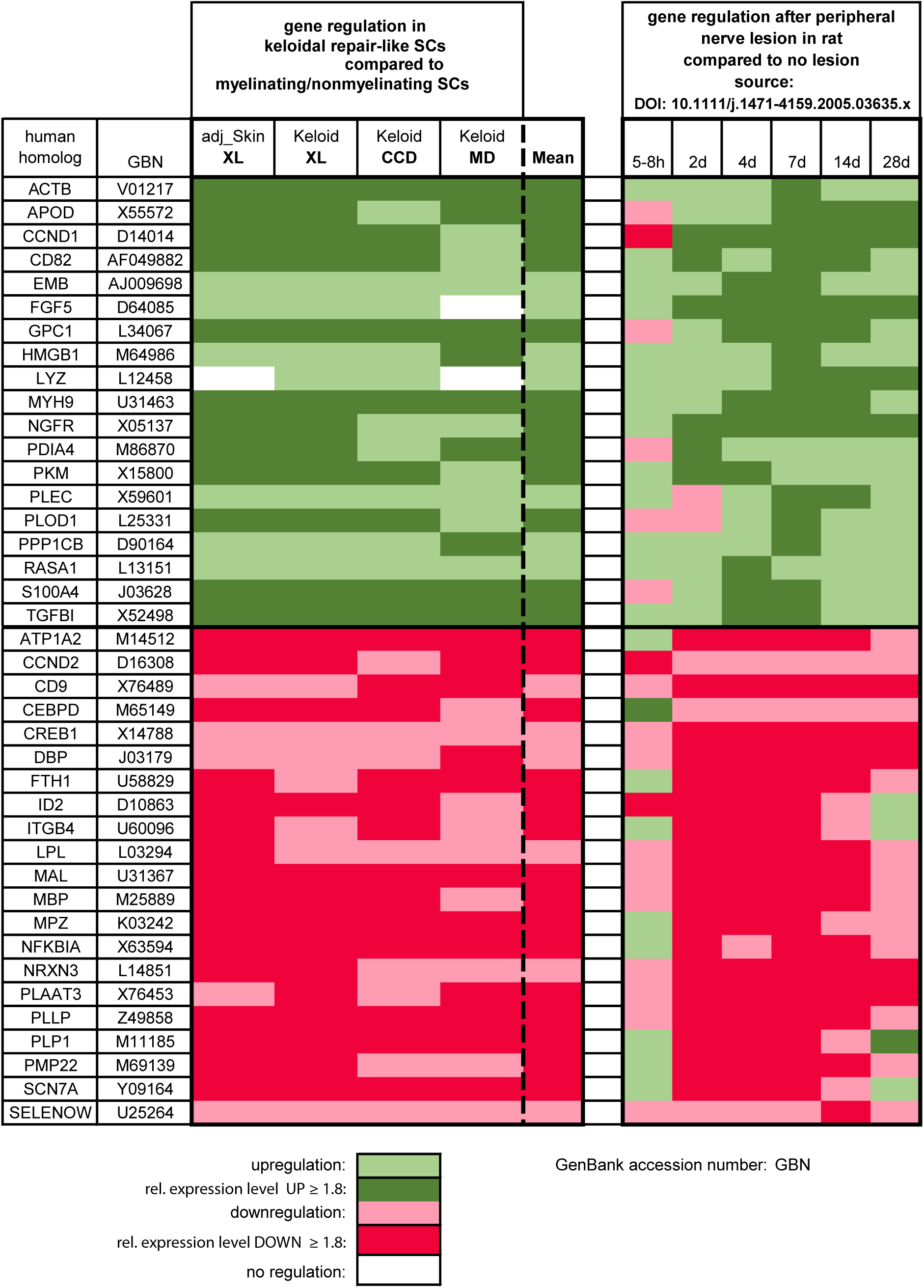
Gene regulation of keloidal Schwann cells matches partially with gene regulation upon neural damage. Heatmap of genetic conformities between keloidal Schwann cells and activated nerve-associated cells upon neuronal damage.

To further identify novel signalling molecules involved in the development of keloidal pro-fibrotic Schwann cell, we performed pseudotime trajectory analysis. Therefore, we re-integrated all individually identified Schwann cell clusters of each study together (Figure 6a). A UMAP, coloured by the previously defined Schwann cell subtypes, showed a clear demarcation of the different Schwann cell populations (Figure 6b). Pseudotime trajectory calculation suggested that myelinating and non-myelinating Schwann cells dedifferentiate towards one common branching point and then further into pro-fibrotic Schwann cells (Figure 6c). Since *JUN, STAT3, ARTN, BDNF, GDNF, SHH* and *OLIG1*, are known decisive factors for the development of repair Schwann cells upon neural damage ^16-19^, we next plotted their expression on the pseudotime trajectory. The transcription factors *JUN* and *STAT3* were highly expressed, specifically in cells at the branching point, confirming their role in the de-differentiation process of repair Schwann cell (Figure 6d). Of note, other described factors involved in repair Schwann cell development in peripheral nerves (*ARTN, BDNF, GDNF, SHH, OLIG1* and *IGFBP2*) were not or only weakly expressed in keloidal Schwann cells (Figure 6d). In addition, our analysis also identified several specific groups of transcription factors significantly enriched at the branching point (Figure 6h), such as members of the AP-1 family (Figure 6e), kruppel like factors (KLF) (Figure 6f) and immediate early response genes (IER) (Figure 6g) as well as some other transcription factors (*ATF3, EGR1, HES1, ZFP36*), suggesting an involvement in keloidal repair-like, pro-fibrotic Schwann cell development.

**Figure 6:**
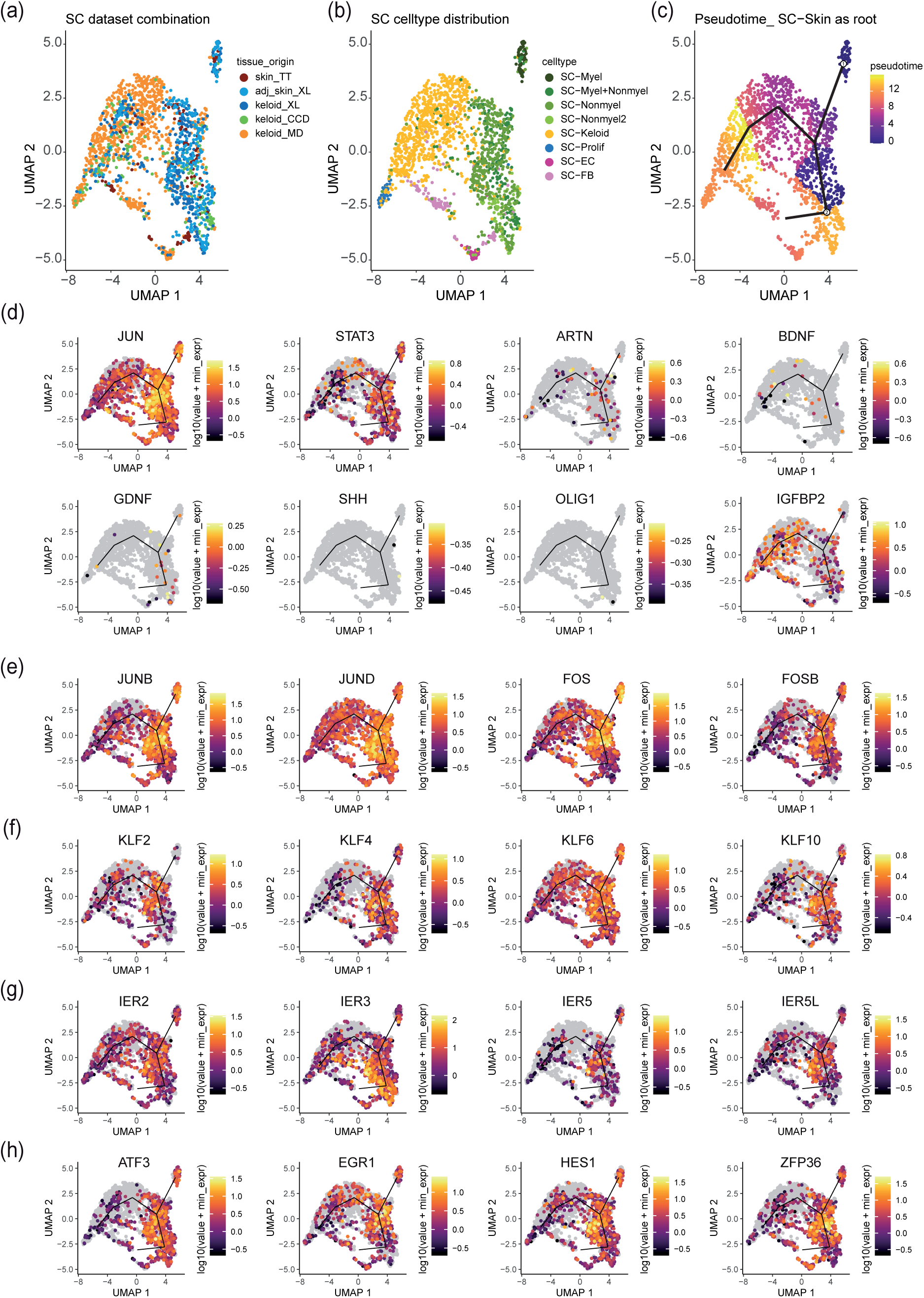
Pseudotime analysis uncovers pivotal genes in the de-differentiation track of Schwann cells. UMAP-Plots combining Schwann cells detected in all individual computations coloured by tissue source (a) and cell type (b) and peudotime trajectory with principal graph (c, myelinating and non-myelinating Schwann cell as root). (d) Feature plots with pseudotime track of Jun proto-oncogene (*JUN*), signal transducer and activator of transcription 3 (*STAT3*), artemin (*ARTN*), brain-derived neurotrophic factor (*BDNF*), glial cell derived neurotrophic factor (*GDNF*), sonic hedgehog (*SHH*), oligodendrocyte transcription factor 1 (*OLIG1*) and insulin like growth factor binding protein 2 (*IGFBP2*). (e) Feature Plots of the activator protein 1 (AP-1) members transcription factor jun-B (*JUNB*), transcription factor jun-D (*JUND*), FOS proto-oncogene (*FOS*) and FOSB proto-oncogene. (f) Feature Plots of kruppel-like factors (KLF) members 4 (*KLF4*), 6 (*KLF6*) and (*KLF10*). (g) Feature Plots of the immediate early response (IER) genes immediate early response 2 (*IER2*), immediate early response 3 (*IER3*), immediate early response 5 (*IER5*) and immediate early response 5 like (*IER5L*). (h) Feature Plots of activating transcription factor 3(*ATF3*), early growth response 1 (*EGR1*), transcription factor hairy and enhyncer of split-1 (*HES1*) and zinc finger protein 36 homolog (*ZFP36*).

## Discussion

Recently, we have identified an important role of Schwann cells in the pathogenesis of keloids ^31^. Using scRNAseq, we were able to demonstrate that a high number of Schwann cells is present in keloids, and that these cells significantly contribute to the over-production of the extracellular matrix ^31^. Interestingly, these Schwann cells were not associated with axons and showed a non-classical repair-like phenotype. However, their exact transcriptional profile and marker genes unambiguously determining this keloidal Schwann cell type have not been described yet. Although scRNAseq represents a powerful tool for studying gene expression in organs and tissues at a single cell resolution, comparability of different scRNAseq data sets and generation of reproducible data are still challenging. Donor variabilities, differences in the tissue dissociation procedure and differences in bioinformatics processing of the data are the main reasons for heterogeneous results ^50-52^. Such methodological differences could significantly affect absolute cell numbers and gene expression patterns. As Schwann cell numbers are already strongly dependent on the body site ^53^ and certain diseases further influence the number and expression profile of Schwann cells in the skin ^31,54^, a combined evaluation of several data sets was essential to decipher the transcriptional profile of the newly identified keloidal Schwann cell type.

So far, several studies have underlined the impact of the tissue dissociation procedure on gene expression ^50-52^. The commercially available whole skin dissociation kit is the most commonly used skin dissociation method for scRNAseq experiments ^36,55^, and was used in 3 of 4 studies analysed (Table 2). Only in one study, keloids were digested with dispase II and collagenase IV for two hours. Interestingly, this data set contained significantly fewer Schwann cells, suggesting that the use of the whole skin dissociation kit yields more reliable data representing all cell types present in the tissue of interest. In the other three studies, the tissue was digested for different time periods. Of note, keloid samples generated by an overnight dissociation step showed significantly more myelinating Schwann cells compared to the studies using a shorter dissociation time (up to 2.5 hours), indicating that the dissociation of Schwann cells attached to axons is difficult and can be improved by a longer digestion period. This finding is in line with established isolation methods of Schwann cells from human peripheral nerves that include an overnight dissociation step (^8,56^). Furthermore, in a recent publication describing a method for the isolation of Schwann cells from healthy skin, the authors discussed pre-culturing steps to disintegrate the nerve for more efficient Schwann cell isolation ^57^.Nevertheless, given the high plasticity of Schwann cells ^2^ and unwanted effects of long lasting isolation protocols on the transcriptome, short term isolation protocols are preferable. In contrast to myelinating and non-myelinating Schwann cells, we found considerable numbers of keloidal pro-fibrotic Schwann cells in all datasets, indicating that Schwann cells that are not associated with an axon can be easily isolated from the tissue.

To compare the phenotype of keloidal Schwann with those of normal scars, we also included scRNAseq data from normal scars in our analyses. Interestingly, only after a comprehensive computation of all datasets together, we were able to identify a few Schwann cells in normal scars. The lack of Schwann cells in normal scars is in line with a recent publication reporting an important role of Schwann cells during wound healing in mice ^21^. Parfejevs *et al*. showed that murine Schwann cells de-differentiation and re-enter the cell-cycle after wounding and promote wound healing by releasing paracrine factors, thereby inducing myofibroblast differentiation ^21^. Importantly, these Schwann cells completely disappeared from the wounded area after successful completion of wound healing ^58^. Whether these Schwann cells convert into other cells, such as fibroblasts, or migrate back to an axon still remains to be elucidated. The low numbers of Schwann cells detected in normal human scars suggest that such a mechanism might also be apparent in human wound healing, and that the development of keloids might be associated with defects in removing these Schwann cells after wound healing. However, further studies are necessary to fully elucidate the impact of Schwann cells on human wound healing.

The adjacent skin around keloids has been reported to exhibit features typical for keloids ^59-68^. Strikingly, our bioinformatics analysis also uncovered keloidal pro-fibrotic Schwann cells in one dataset of the keloid-adjacent skin. This finding supports our assumption that the persistence of keloidal Schwann cells in the dermis contributes to disease progression. Two scenarios by which these Schwann cells might affect keloid progression appear reasonable. Firstly, keloidal Schwann cells spread into the surrounding healthy skin, generating a milieu that favours the growth of keloids. Secondly, the continuously growing keloids might affect nerves in the surrounding healthy skin and trigger de-differentiation of Schwann cells, which in turn further contribute to ECM over-production as reported previously ^31^. Our transcriptomics data and bioinformatics analyses has built a good basis for further studies addressing these open questions in more sophisticated *in vivo* studies.

Injury of peripheral nerves causes de-differentiation of both types of mature Schwann cells (myelinating and non-myelinating) into repair Schwann cells ^69-71^. Our pseudotime trajectory calculations suggest that myelinating and non-myelinating Schwann cells of the skin de-differentiate into pro-fibrotic, repair-like Schwann cells in keloids. Interestingly, many of the marker genes specifically expressed by *bona* fide repair Schwann cells (*STAT3, ARTN, BDNF, GDNF, SHH, OLIG1 and IGFBP2*) were not or only weakly detected in keloidal Schwann cells. However, c-Jun, a key factor for guiding de-differentiation and enabling proper function of repair Schwann cells ^16,70,72-75^, was also strongly expressed in keloidal Schwann cells, especially at the branching point of the pseudotime trajectory. However, in fully de-differentiated keloidal Schwann cells, c-Jun expression levels were again decreased. Interestingly, our analyses revealed that several other transcription factors, including other AP-1 members (*JUNB, JUND, FOS, FOSB*), members of the kruppel-like factor family (*KLF2, KLF4, KLF6, KLF10*) and members of the immediate early response gene family (*IRE2, IRE3, IER5, IER5L*) were similarly regulated, suggesting that these transcription factors might also play an important role in the de-differentiation process of skin Schwann cells. Whereas the function of most of these factors in Schwann cells needs further in-depth investigations, several members of the KLF family have already been reported to be crucial for Schwann cell differentiation ^76-80^. Especially the high expression of KLF4 suggest that keloidal Schwann cells indeed underwent a de-differentiation process. The reason for the lack of *bona fide* repair Schwann cell markers in keloidal Schwann cells is currently not known. As c-Jun is the major regulator of most of these marker genes ^16^, its down-regulation might be responsible for this observation. Although c-Jun activation is important for the induction of repair Schwann cell development after neural damage, chronic denervation was shown to strongly reduce c-Jun levels in Schwann cells ^9^. We therefore hypothesize that due to the prolonged axon-free occurrence of Schwann cells in keloids, their expression pattern changes significantly. Whether these keloidal Schwann cells, despite the loss of expression of several repair marker genes, still represent functional repair Schwann cells, has to be determined in future experiments. However, the comparison of our dataset with a dataset of an acute nerve lesion ^15^ revealed that a high number of repair genes was similarly regulated in both Schwann cell types, suggesting that at least some parts of the repair Schwann cell function are still conserved in keloidal Schwann cells.

As we have previously shown that keloidal Schwann cells significantly contribute to ECM formation ^31^, it is conceivable that keloidal Schwann cells acquired pro-fibrotic properties. This hypothesis is supported by the identified gene set that contains several pro-fibrotic genes (*COL1A1, COL7A1, ELN, IGFBP3, IGFBP5, ITGB1, LOXL2, MMP15, S100A16, SPARCL1, TAGLN, TGFBI, TPM2*). The identification of this specific gene set enables further investigations on the contribution of Schwann cells to fibrosis in other fibrotic disorders, such as liver cirrhosis, idiopathic pulmonary fibrosis or renal fibrosis. Considering the current literature and our new data, we hypothesize that there are at least two types of repair(-like) Schwann cells. One type occurring temporarily after acute injury to functionally regenerate a disrupted nerve, and another type persisting after completed tissue regeneration in the wounded area for a long time period, thereby acquiring pro-fibrotic properties ^9,21^. Why this Schwann cell type is not removed after wound healing is currently not known and matter of ongoing experiments.

In conclusion, we confirmed the presence of a repair-like, pro-fibrotic Schwann cell type in keloids from three independent scRNAseq data sets and identified a conserved expression pattern of twenty-one genes that specifically characterize this Schwann cell type. This special repair-like, pro-fibrotic Schwann cell type present in keloids exhibits distinct genetic differences compared to classical repair Schwann cells but presumably arise from a common initial injury event. Our study has built the foundation for further studies to investigate the frequency of these cells and the contribution to fibrotic processes in other organs. With respect to the described bioinformatics analyses, our study provides a practical strategy to reliably analyse even small cell clusters and obviate wrong cluster assignment.

## Data Availability

All data produced are available online at

https://www.ncbi.nlm.nih.gov/geo/query/acc.cgi

https://www.ncbi.nlm.nih.gov/geo/query/acc.cgi

https://dom.pitt.edu/wp-content/uploads/2018/10/Skin_6Control_rawUMI.zip

https://dom.pitt.edu/wp-content/uploads/2018/10/Skin_6Control_Metadata.zip

https://ngdc.cncb.ac.cn/gsa-human/browse/HRA000425

## Data Availability Statement

All ScRNAseq data are publicly accessible in NCBI’s Gene Expression Omnibus (GSE181316, GSE163973), at the Genome Sequencing Archive (BioProject PRJCA003143) and under the following links: https://dom.pitt.edu/wp-content/uploads/2018/10/Skin_6Control_rawUMI.zip https://dom.pitt.edu/wp-content/uploads/2018/10/Skin_6Control_Metadata.zip

## Acknowledgments

The authors would like to thank HP Haselsteiner and the CRISCAR Familienstiftung for their ongoing support of the Medical University/Aposcience AG public private partnership aiming at augmenting basic and translational clinical research in Austria/Europe. We acknowledge the core facilities of the Medical University of Vienna, a member of Vienna Life Science Instruments. The study was funded by the FFG Grant “APOSEC” (852748 and 862068; 2015-2019), the Vienna Business Agency “APOSEC to clinic” (ID 2343727, 2018-2020) and by the Aposcience AG under the direction of group leader HJA. MM was funded by the Sparkling Science Program of the Austrian Federal Ministry of Education, Science and Research (SPA06/055).

## Author contributions

Conceptualization, M.D., T.W. and M.M.; methodology, M.D., M.W., D.C. and M.M.; software, M.D., M.W., D.C., and K.K.; validation, M.D. and M.M.; formal analysis, M.D.; investigation, M.D. and D.C.; resources, H.J.A. and M.M.; data curation, M.D.; writing original draft preparation, M.D., M.L. and M.M.; writing, review and editing, M.D., M.W., T.W., M.L., K.K., D.B., V.V. and M.M.; visualization, M.D.; supervision, M.M.; project administration, M.D. and M.M.; funding acquisition, M.M. and H.J.A. All authors have read and agreed to the final version of the manuscript.

## Institutional Review Board Statement

The study was conducted according to the guidelines of the Declaration of Helsinki. The use of resected skin and keloids has been approved by the ethics committee of the Medical University of Vienna (votes 217/2010 and 1190/2020).

## Informed Consent Statement

Written informed consent was obtained from all donors

## Supplementary Figures

**Figure S1:**
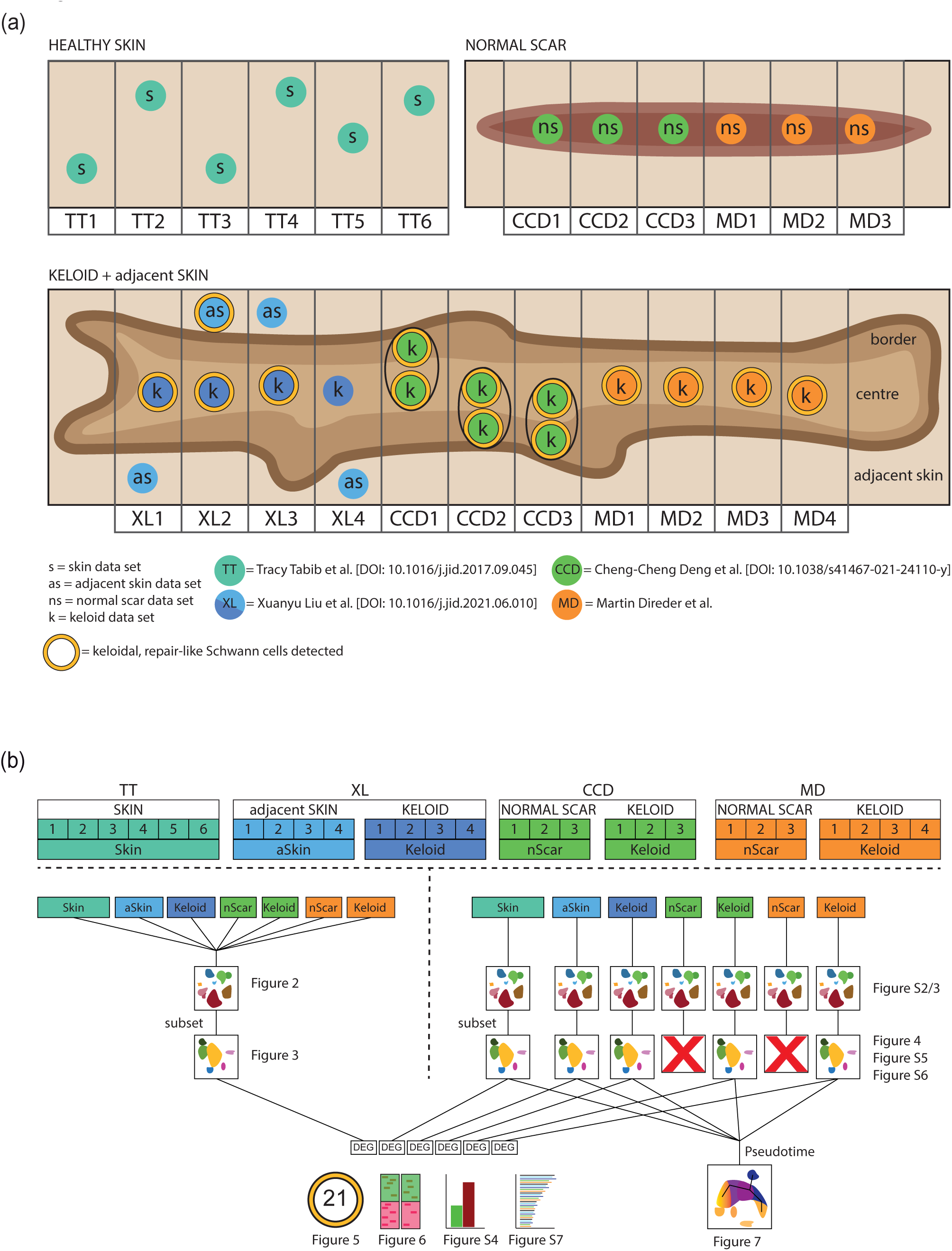
Graphical scheme of the included datasets and bioinformatics method overview. (a) Schematic illustration of the regional sampling point of every dataset. (b) Synoptical chart of all dataset combinations included in this study with link for each respective figure. skin (s), adjacent skin (as/aSkin), normal scar (ns/nScar), keloid (k); data source: Tracy Tabib et al., 2018 (TT); Xuanyu Liu et al., 2021 (XL), Cheng-Cheng Deng et al., 2021 (CCD), Martin Direder et al., 2022 (MD); DEG= differentially expressed genes;

**Figure S2:**
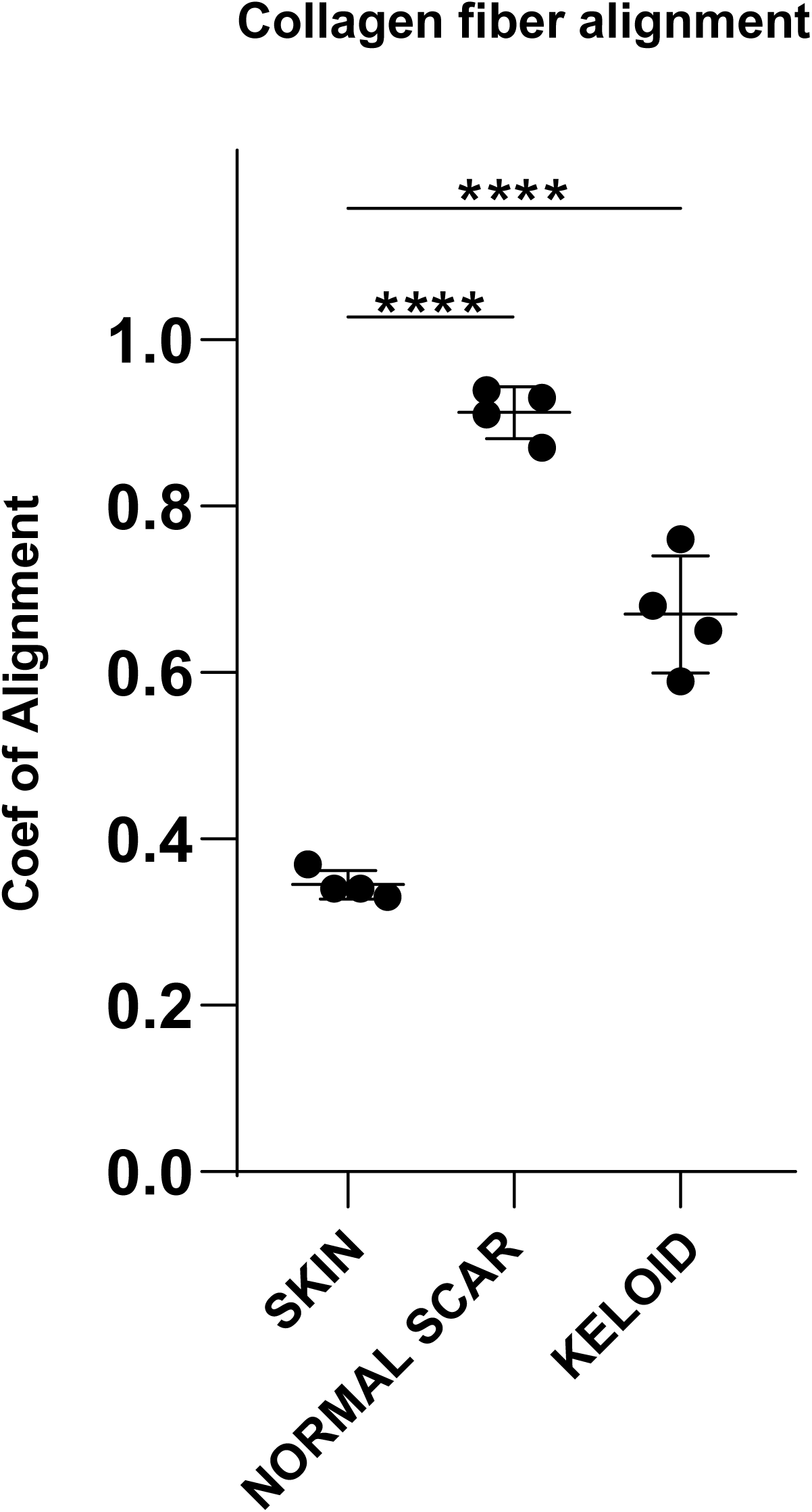
Collagen alignment in skin, normal scar and keloid. Evaluation of the alignment coefficient of healthy skin, normal scar and keloid tissue. Maximum and minimum value with <1.5 interquartile range is shown by whiskers. The middle line represents the mean. Normally distributed data were compared by one-way ANOVA with Tukey *post hoc* test. ****p <0.0001.

**Figure S3:**
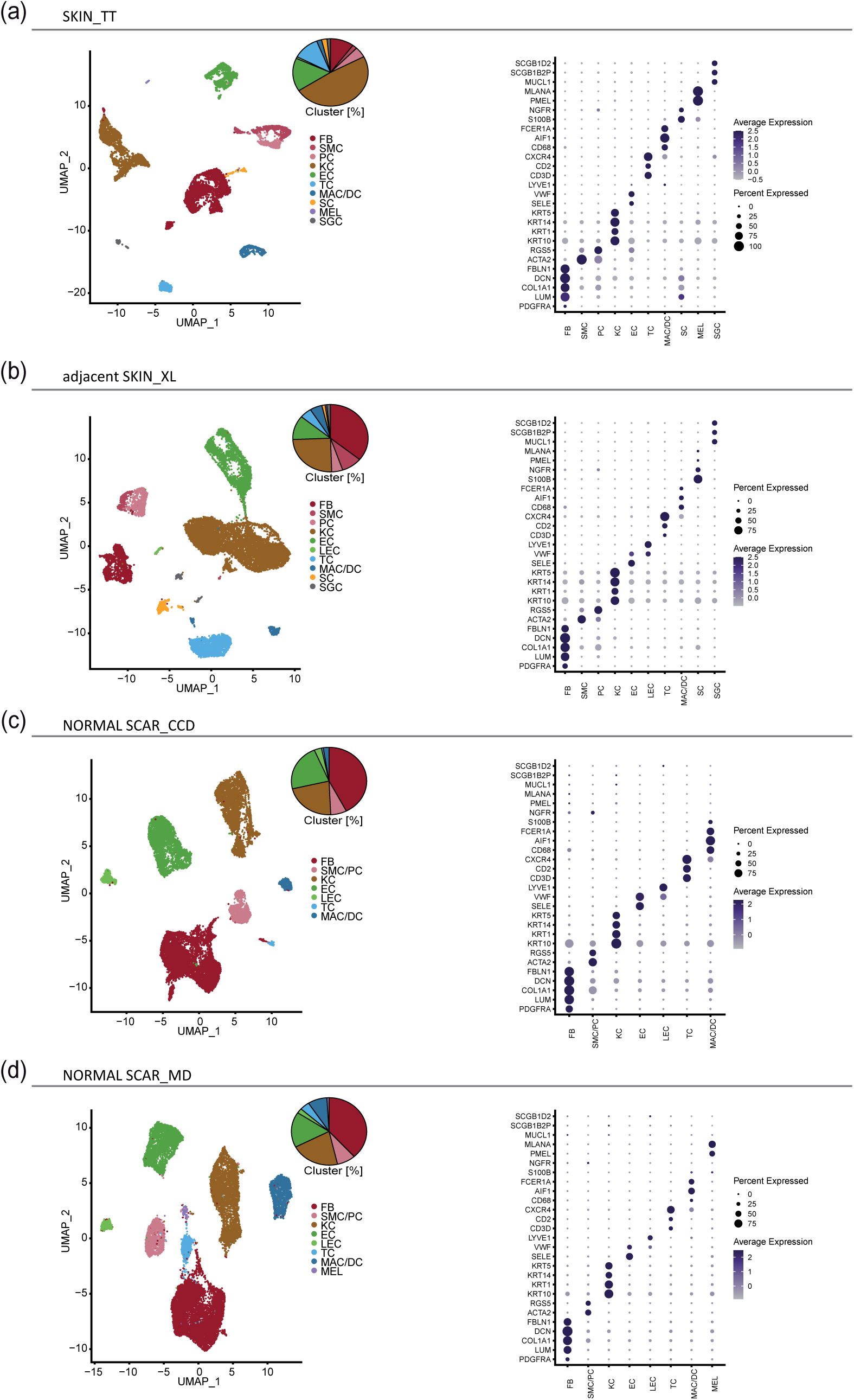
Individual dataset integration unravels cluster consistency. UMAP-Plots and Dot plots individual integrations of datasets from distinct conditions and sources: SKIN_TT, n=6 (a); adjacent SKIN_XL, n=4 (b); NORMAL SCAR_CCD, n=3 (c); NORMAL SCAR_MD, n=3 (d); cluster classification as fibroblasts (FB), smooth muscle cells (SMC), pericytes (PC), smooth muscle cells and pericytes (SMC/PC), keratinocytes (KC), endothelial cells (EC), lymphatic endothelial cells (LEC), T-cells (TC), macrophages (MAC), dendritic cells (DC), macrophages and dendritic cells (MAC/DC), Schwann cells (SC), melanocytes (MEL), erythrocytes (ERY) and sweat gland cells (SGC); Pie plots show cluster composition within each dataset combination. Dot plots showing well-known marker genes to characterise clusters: platelet-derived growth factor receptor A (*PDGFRA*), lumican (*LUM*), collagen type I alpha 1 (*COL1A1*), decorin (*DCN*), fibulin-1 (*FBLN1*) for FB, actin alpha 2 (*ACTA2*), regulator of G protein signalling 5 (*RGS5*) for SMC and PC, keratin 10 (*KRT10*), keratin 1 (*KRT1*), keratin 14 (*KRT14*), keratin 5 (*KRT5*) for KC, E-selectin (*SELE*), von willebrand factor (*VWF*) for EC, lymphatic vessel endothelial hyaluronan receptor 1 (*LYVE1*) for LEC, *CD3D*, cluster of differentiation 2 (*CD2*), c-x-c chemokine receptor type 4 (*CXCR4*) for TC, cluster of differentiation 68 (*CD68*), allograft inflammatory factor 1 (*AIF1*) for MAC, Fc fragment of IgE receptor Ia (*FCER1A*) for DC, S100 calcium binding protein B (*S100B*), nerve growth factor receptor (*NGFR*) for SC, premelanosome protein (*PMEL*), melan-a (*MLANA*) for MEL; Dot size symbolizes percentage of cells expressing the gene, colour gradient shows level of average gene expression.

**Figure S4:**
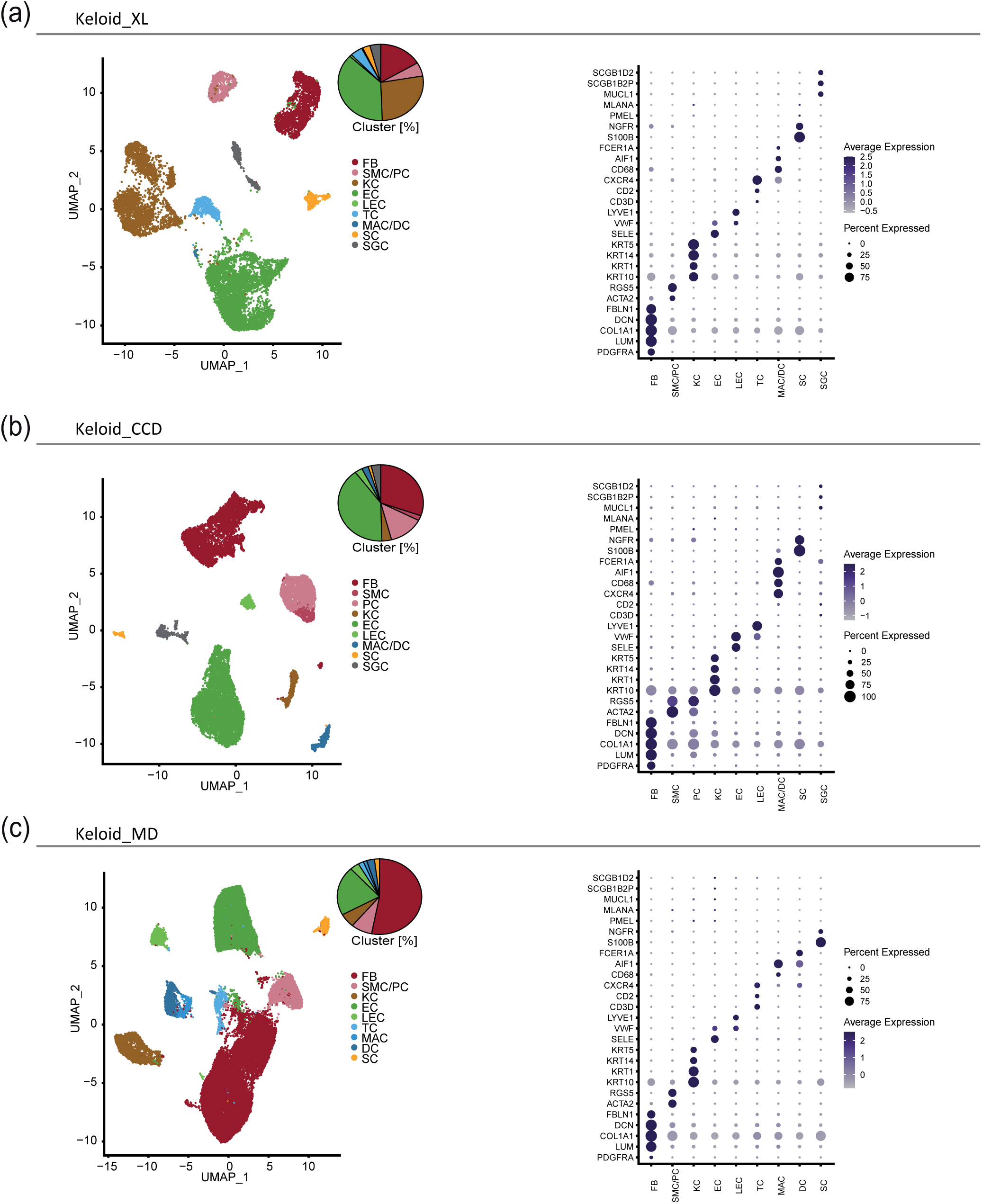
Individual dataset integration unravels cluster consistency. UMAP-plots and DotPlots individual integrations of datasets from distinct conditions and sources: KELOID_XL, n=4 (a); KELOID_CCD, n=3 (b); KELOID_MD, n=4 (c); cluster classification as fibroblasts (FB), smooth muscle cells (SMC), pericytes (PC), smooth muscle cells and pericytes (SMC/PC), keratinocytes (KC), endothelial cells (EC), lymphatic endothelial cells (LEC), T-cells (TC), macrophages (MAC), dendritic cells (DC), macrophages and dendritic cells (MAC/DC), Schwann cells (SC), melanocytes (MEL), erythrocytes (ERY) and sweat gland cells (SGC); Pie plots show cluster composition within each dataset combination. Dot plots showing well-known marker genes to characterise clusters: platelet-derived growth factor receptor A (*PDGFRA*), lumican (*LUM*), collagen type I alpha 1 (*COL1A1*), decorin (*DCN*), fibulin-1 (*FBLN1*) for FB, actin alpha 2 (*ACTA2*), regulator of G protein signalling 5 (*RGS5*) for SMC and PC, keratin 10 (*KRT10*), keratin 1 (*KRT1*), keratin 14 (*KRT14*), keratin 5 (*KRT5*) for KC, E-selectin (*SELE*), von willebrand factor (*VWF*) for EC, lymphatic vessel endothelial hyaluronan receptor 1 (*LYVE1*) for LEC, *CD3D*, cluster of differentiation 2 (*CD2*), c-x-c chemokine receptor type 4 (*CXCR4*) for TC, cluster of differentiation 68 (*CD68*), allograft inflammatory factor 1 (*AIF1*) for MAC, Fc fragment of IgE receptor Ia (*FCER1A*) for DC, S100 calcium binding protein B (*S100B*), nerve growth factor receptor (*NGFR*) for SC, premelanosome protein (*PMEL*), melan-a (*MLANA*) for MEL; Dot size symbolizes percentage of cells expressing the gene, colour gradient shows level of average gene expression.

**Figure S5:**
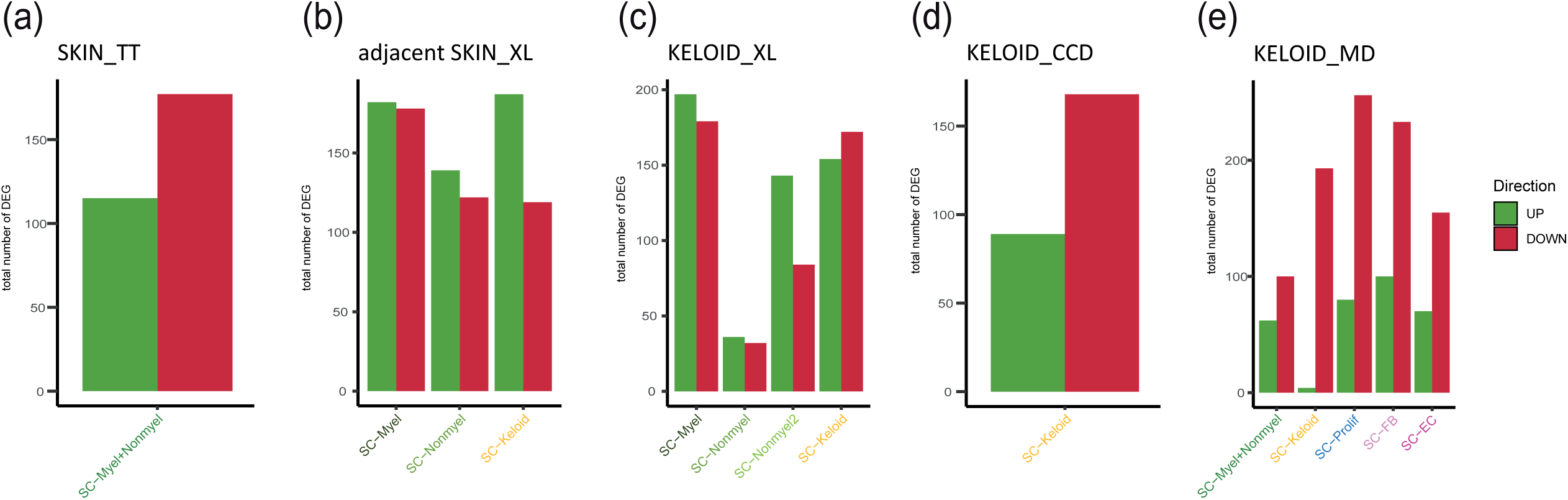
Total number of differentially expressed genes in each individual computation. Bar plots show the total number of differentially expressed genes (fold change ≥2) between all Schwann cell cluster of SKIN_TT (a); adjacent SKIN_XL (b); KELOID_XL (c); KELOID_CCD (d); KELOID_MD (e); green= UP regulation, red= DOWN regulation

**Figure S6:**
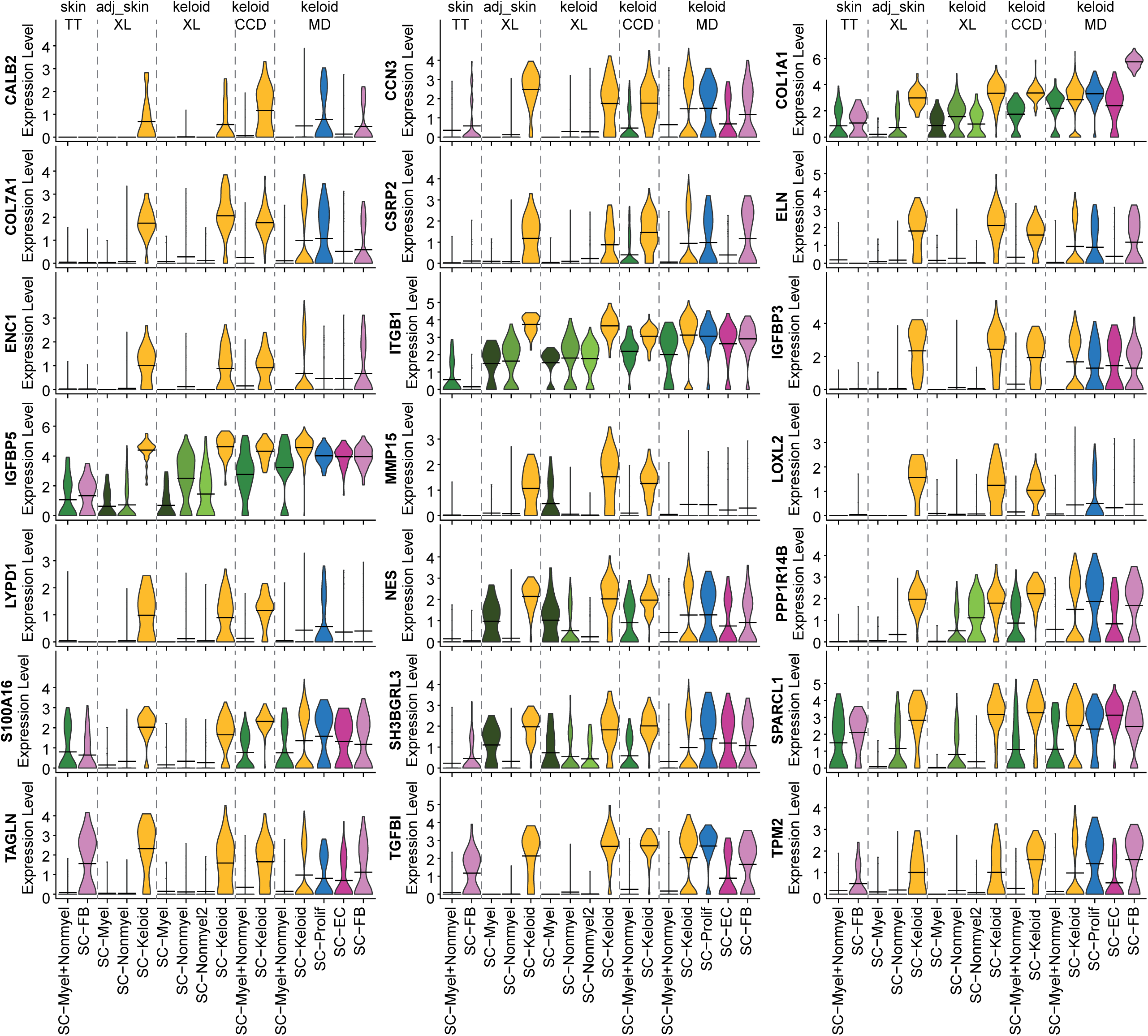
Extracellular matrix associated gene expression in SC-Keloid of distinct sources. Violin plots show the expression of the identified SC-Keloid expression pattern in all Schwann cell subtypes. Crossbeams mark the mean expression, vertical lines show maximum expression, violin width symbolizes frequency of cells on a distinct expression level.

**Figure S7:**
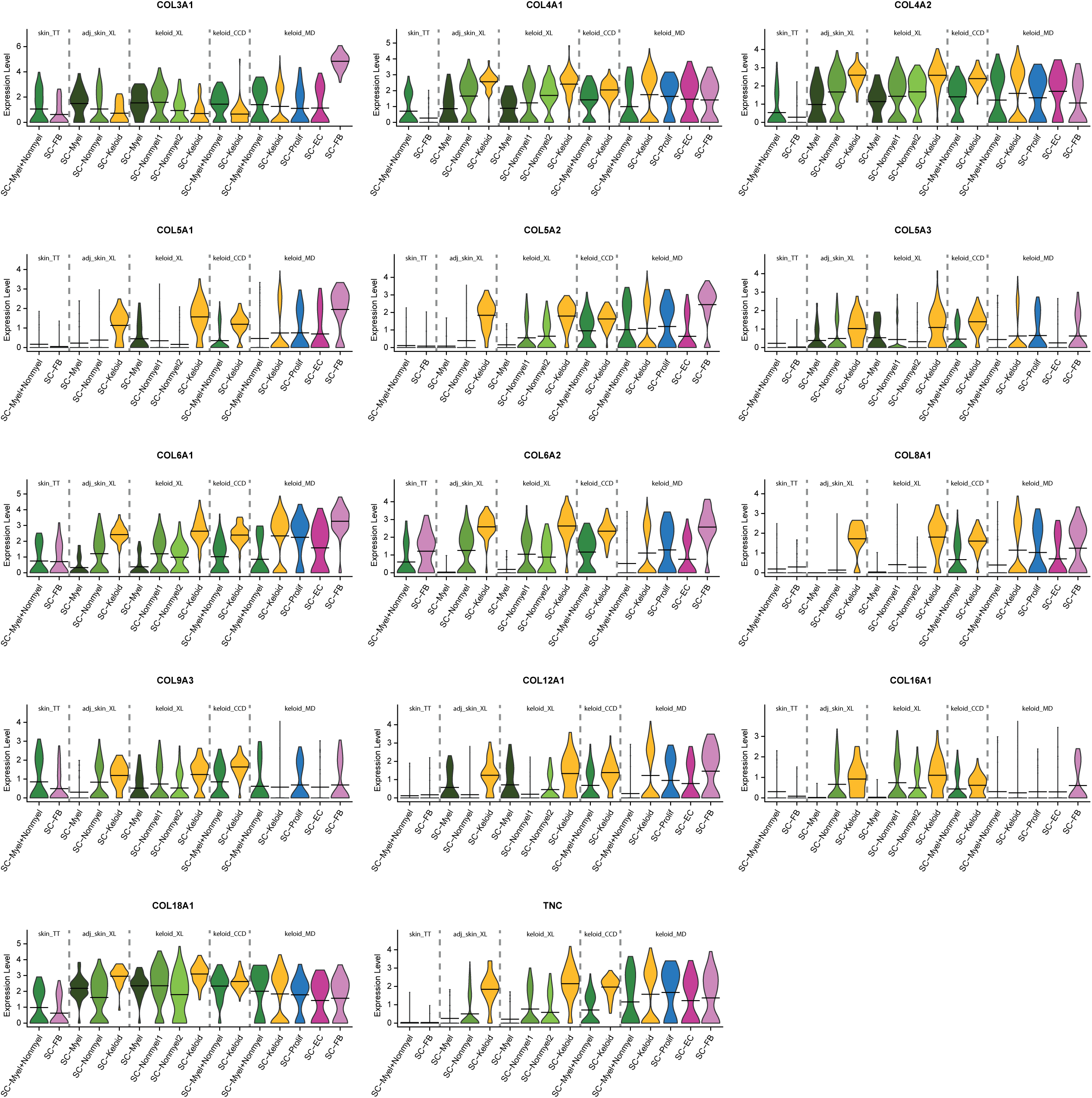
Extracellular matrix associated gene expression in SC-Keloid of distinct sources. Violin plots show the expression of several genes associated with matrix formation in all Schwann cell subtypes. Crossbeams mark the mean expression, vertical lines show maximum expression, violin width symbolizes frequency of cells on a distinct expression level.

**Figure S8:**
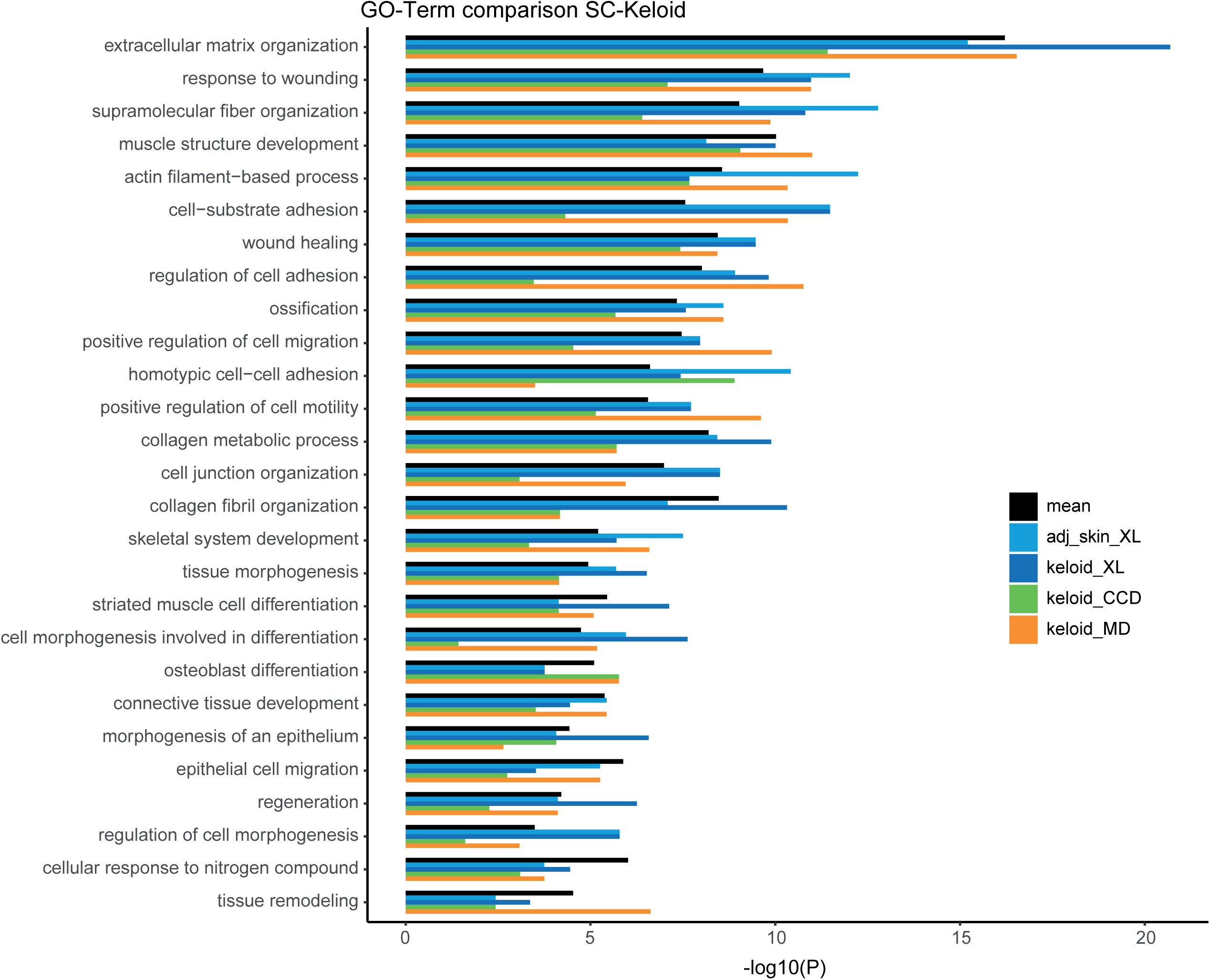
Corresponding GO-Terms encourage reported functions of keloidal Schwann cells. Enrichment analysis of top 100 upregulated genes resulted by data confrontation of SC-Keloid with myelination and nonmyelinating Schwann cells. Bar length shows statistical significance; mean value is depicted in black.

